# Agricultural Work, Malaria Prevalence, and Mediating Factors: A Cross-Sectional Analysis of Data from 15 Sub-Saharan African Countries to Inform Risk Stratification and Intervention Planning

**DOI:** 10.1101/2025.07.21.25331926

**Authors:** Ifeoma D. Ozodiegwu, Grace Legris, Chilochibi Chiziba, Laurette Mhlanga

## Abstract

**Background:** Although linkages between agricultural work and malaria have been widely studied, country-specific information on disease burden, intervention use, and the pathways through which adult agricultural work affects the health of children in the same household remains limited. This study examines the relationship between agricultural work, malaria infection, and related factors among children under five, highlighting subnational variation across urban and rural areas within each country.

**Methods:** Demographic and Health Survey data from 15 sub-Saharan African countries were analyzed to generate descriptive statistics on malaria prevalence, bed net use, and household, age-related, and environmental characteristics of sampled children. Potential mediators were identified by examining their associations with both self-reported agricultural occupation of an adult household member and malaria positivity. Mediation analysis was conducted using logistic regression models within a Structural Equation Modeling framework to estimate indirect, direct, and total effects. The percentage mediated was calculated using percentile bootstrapping.

**Results:** Children in agricultural households face a higher risk of malaria than those in non-agricultural households, despite similar rates of bed net use. Substantial country-level variation in malaria prevalence and net use underscores the complex interplay between agricultural occupation, intervention uptake, and malaria risk. Although less common overall, urban agricultural households remain concentrated in populous regions of several countries. Mediation analysis identified wealth, enhanced vegetation index, treatment-seeking behavior, and housing quality as the strongest mediators of this relationship in both urban and rural settings.

**Conclusions:** These findings offer actionable insights for countries aiming to better target malaria interventions, particularly in urban areas. The varied mediating factors highlight the need for an integrated approach that addresses housing quality, environmental risk, treatment-seeking behavior, and socioeconomic conditions to protect young children in agricultural households and strengthen malaria control efforts amid shrinking resources.

## Background

Since 2000, the global fight against malaria has seen substantial progress, with marked declines in malaria-related deaths [1]. These gains are largely credited to improved housing and infrastructure [2–4], as well as the widespread adoption of effective prevention and treatment tools, particularly long-lasting insecticidal nets (LLINs) and artemisinin-based combination therapies [5], across sub-Saharan Africa, where over 90% of global malaria cases and deaths have occurred in the past decade [1].

However, this progress is now under threat due to disruptions to health systems from COVID-19 [6,7], emerging infectious diseases [8,9], conflict [10], and climate-related disasters [11,12], which are contributing to a concerning reversal of gains. Recent estimates show an upward trend in malaria cases and deaths [1]. This situation is further exacerbated by recent United States government cuts to global health funding. As the largest international donor supporting malaria control efforts, the U.S. funds critical interventions—including the distribution of LLINs, diagnostic tools, and treatments—that prevent millions of cases and save lives annually [13]. These funding reductions may jeopardize essential program infrastructure and accelerate the resurgence of malaria, particularly in high-burden settings.

In this context, it is increasingly urgent for countries to optimize the use of limited resources by prioritizing populations at greatest risk. Agricultural communities in sub-Saharan Africa represent one such high-risk group. Several factors elevate malaria risk in these settings. Agricultural households often live and work in environments, such as irrigated rice fields, rainfed croplands, and forested areas, that provide ideal breeding conditions for efficient malaria vectors [14–17]. Additionally, agricultural practices like introducing new crop species or employing slash-and-burn techniques can transform otherwise unsuitable areas into vector habitats, thereby increasing human exposure and entomological inoculation rates [18].

Moreover, the intensive use of agricultural pesticides, many of which share active ingredients with those used in vector control, can reduce natural predators and foster insecticide resistance [18,19]. This resistance undermines the efficacy of key interventions such as LLINs and indoor residual spraying, which predominantly target indoor-biting mosquitoes. As sub-Saharan African countries urbanize and transition from agriculture-based to service-based economies, urban communities engaged in agricultural work may serve as reservoirs of infection, contributing to a heterogeneous malaria burden.

Children under five in agricultural households may face heightened vulnerability to malaria due to biological susceptibility and the heighted malaria risk in their communities. Yet, there remains a paucity of generalizable, country- and subnational-level evidence quantifying this risk. Generating country-specific data comparing malaria burden between children in agricultural versus non-agricultural households—while accounting for factors like LLIN use, where national malaria programs and their partners are making significant investments [20]— can support more targeted resource allocation and intersectoral collaboration. Understanding the geographic distribution of agricultural employment can further inform the tailoring of malaria interventions to areas of highest need.

In this article, we analyze household survey data to explore differences and trends in malaria prevalence and bed net use among children under five, stratified by whether a resident adult works in agriculture or another sector, and disaggregated by country and urban–rural residence. We map subnational variation in the concentration of agricultural households in urban areas and describe household, environmental, and individual characteristics of children based on the occupation of resident adults. Finally, we estimate the extent to which various factors mediate the relationship between resident adult occupation and childhood malaria risk.

## Methods

### Data

We used the *rdhs* package in R to query the Demographic and Health Survey (DHS) API and retrieved survey datasets from 2010 onwards that included malaria test results and occupation information. The DHS are nationally representative, multistage cluster-level surveys designed by the DHS Program to provide reliable data on health and population in developing countries [21]. In these surveys, interviews were conducted with a random sample of households in each selected cluster. In each household, all women aged 15–49 years, and a subsample of men in the same age range respond to questions on behalf of themselves and their household members. Children aged 6–59 months in the sampled households were tested for malaria using rapid diagnostic tests (RDT) and/or microscopy [21]. We excluded countries with malaria test positivity rates of ≤1% due to concerns about statistical power to detect malaria prevalence differences in children under the age of five group by the occupation of resident adults. The two most recent surveys with malaria test positivity rates >1% were retained for each country, resulting in one dataset for eight countries and two datasets (one from the most recent year and one from the preceding year) for seven countries. Surveys were conducted between 2012 and 2021 across the 15 African countries.

The analysis dataset included environmental factors — temperature, precipitation, relative humidity, and enhanced vegetation index (EVI) — extracted at the cluster-level using buffers with a radius of 2000 meters, to account for the DHS displacement designed to protect participant confidentiality [22]. These factors were chosen because they influence malaria transmission by affecting the duration and geographic extent over which malaria parasites and Anopheles vectors can coexist and reproduce [23–25]. Key demographic factors known to influence malaria risk, including household size [26,27], gender [28–30], and socioeconomic status [30,31] were retained in the dataset. Net use, medical treatment-seeking for fever symptoms, used as a proxy indicator for malaria treatment seeking behaviors, and stunting were also among the factors retained in the dataset.

Values of all covariates not directly obtained from the DHS (e.g. environmental or satellite-derived variables) were matched at the cluster-level to the specific year in which the DHS survey was conducted in each country, to ensure that covariate timing corresponded to the survey period. This meant that each child in the dataset inherited the cluster value for these covariates. A detailed description of all considered covariates and their sources can be found in Table 1.

**Table 1.** Variable names, definitions, and sources for variables considered for modeling.

| Variable Name | Variable Definition | Data Source |
| --- | --- | --- |
| Dependent variable |  |  |
| Malaria test result | Whether a child aged 6–59 months tested positive for malaria by microscopy or RDT. | DHS[21] |
| Independent variable |  |  |
| Home type | Whether a household includes an adult who worked in agriculture in the last year. | DHS[21] |
| Covariates |  |  |
| Household size | Number of individuals living in the household where a child aged 6–59 months resided. | DHS[21] |
| Enhanced vegetation index | A satellite-derived measure of vegetation greenness, derived at a 5km resolution, based on near-infrared, red, and blue spectral bands and estimated at a cluster level. Values range from –1.0 to 1.0, with higher values indicating denser or healthier vegetation. Derived from MODIS/006/MOD09GA surface reflectance composites. For methodology, see Huete et al. (2002).[35] | Malaria Atlas Project[36] |
| Precipitation | Estimated total rainfall (in millimeters) for the two months preceding the survey month and year, derived at a 5km resolution from satellite imagery and supplemented with in-situ weather station data. Values are calculated at the cluster level. | Climate Hazards Group InfraRed Precipitation with Station data[37] |
| Relative humidity | The percentage of water vapor pressure relative to air saturation at the survey location on a 27km grid, averaged over the two months preceding the survey month and year, calculated at the cluster level. Saturation is calculated over water for temperatures above 0°C, over ice below –23°C, and interpolated between –23°C and 0°C. | ERA5[38] |
| Temperature | Estimated air temperature (°C) at 2 meters above ground level over a 27km grid, averaged over the two months preceding the survey month and year at a cluster level. | ERA5[38] |
| Age | Child's age in months. | DHS[21] |
| Housing quality | Whether a housing unit where a child aged 6–59 months resided was made of good quality materials such as cement, bricks, or covered adobe. | DHS[21] |
| Wealth | A composite measure of a household’s living standard, based on assets, housing materials, and access to water and sanitation. Households were ranked on a continuous scale using principal components analysis and divided into five wealth quintiles (Richest, Rich, Middle, Poor and Poorest). | DHS[21] |
| Gender | Child’s gender | DHS[21] |
| Severe stunting | Whether a child aged 6–59 months has a height-for-age z-score more than 3 standard deviations below the median of WHO Child Growth Standards. | DHS[21] |
| Net use among children under the age of five years | Whether a child aged 6–59 months slept under an insecticide-treated net (ITN) the night before the survey. | DHS[21] |
| Treatment seeking for fever | A binary variable coded as 1 if the individual lives in a cluster where either (a) no one reported having fever in two weeks prior to the survey (underlying assumption is that treatment seeking is high), or (b) at least 60% of those who reported fever sought medical treatment. It is coded as 0 if fewer than 60% of individuals with fever in the cluster sought treatment. | DHS[21] |

To visualize the geographic distribution of agricultural households, we used shapefiles of first-level administrative subdivisions (e.g. states, regions) for each of the 15 countries included in the analysis. These shapefiles were primarily obtained from the Database of Global Administrative Areas (GADM) [32]. However, because GADM lacked shapefiles for Uganda’s regions, we sourced them from The Humanitarian Data Exchange [33]. Population estimates for 2020 at a 100m resolution were obtained from WorldPop [34] and overlaid with first-level administrative subdivision shapefiles to calculate total population per subdivision in each of the 15 countries.

### Descriptive Analysis

To assess the sample sizes by country, we employed flipped bar plots to provide an overview of the survey-weighted count and percentage of children aged 6–59 months who underwent malaria testing using RDT or microscopy, stratified by their urban or rural residence.

We estimated and visualized the proportion of children testing positive for malaria (via RDT or microscopy) in both agricultural and non-agricultural worker households, overall, by place of residence (urban vs. rural), and by country with an urban-rural breakdown. Similarly, we estimated net use overall, as well as by country and place of residence, for both agricultural and non-agricultural worker households, then computed and visualized the disparities in net use and test positivity rates between these two home types in urban and rural areas. Subsequently, we fit a linear regression model to understand the relationship between disparities in net use and test positivity rates for all countries. For seven countries with two survey datasets containing both malaria results and occupation data, we computed and plotted changes in malaria positivity and net use between the most recent and preceding surveys for both home types. To further explore trends in malaria transmission prevention, we enhanced these line plots by layering an area plot illustrating the annual percentage change in nets distributed, calculated relative to the first year with available data in each country. Data used in computing the annual percentage change in nets distributed was sourced from 2023 World Malaria Report [39] and through email communication with staff of the World Health Organization.

Additionally, we created a series of maps depicting the proportion of agricultural households across each first-level administrative subdivision within each country and highlighting the three most populous first-level subdivisions. Top-down unconstrained population estimates from WorldPop.org [34] were used, as constrained datasets did not provide annual estimates. Unconstrained data were available for 2000–2020, and for survey datasets spanning two years (e.g., Angola 2015–16), population estimates from the latter year were applied. For surveys conducted after 2020, such as Ghana 2022–23, estimates from 2020 were used. Population estimates at a 100m resolution were extracted for the 15 countries and overlaid with first-level administrative subdivision shapefiles to calculate total population per subdivision.

Environmental, household and child age-related characteristics of agricultural and non-agricultural worker households were visualized using boxplots and bar plots.

### Statistical Analysis

#### Mediation Model Development and Implementation

To assess the relationship between home type (agricultural or non-agricultural) and malaria positivity among children under five, we conducted mediation analyses to identify key factors that explain this association. The mediation model aimed to estimate the extent to which the effect of home type on malaria positivity is mediated through household and individual-level variables.

We began by conducting a series of logistic regression models to identify potential mediators. These models assessed whether home type was associated with each covariate and whether each covariate was independently associated with malaria test positivity in the pooled dataset across all countries. To account for potential confounding, logistic regression models assessing the association between each covariate and malaria positivity also adjusted for home type, ensuring that observed associations were not solely attributable to differences in household occupation. Analyses accounted for the DHS’s complex, multistage sampling design using survey weights, implemented via the *survey* package in R. Odds ratios were estimated separately by urban and rural places of residence. A generalized version of the logistic regression model equation is written as follows:

To be included as potential mediators, variables had to meet both theoretical and empirical criteria. Empirically, each variable needed to show a significant association with both malaria positivity and with home type (agricultural vs. non-agricultural) in at least one setting—urban or rural—as shown in Supplementary Tables 2–3. Variables such as child’s age, gender, temperature, relative humidity, and precipitation were excluded as potential mediators in the logistic regression models due to lack of temporal precedence or implausibility of a direct causal pathway from home type.

We used a structural equation modeling framework to estimate the indirect effect (the pathway from home type to malaria positivity through mediators, refer to Fig 1), direct effect (home type to malaria positivity independent of mediators), and total effect (sum of direct and indirect effects). In logistic regression models, wealth was treated as a categorical variable, with each quintile compared to the lowest (Q1) as the reference. In contrast, for mediation analysis, wealth was modeled as an ordinal variable to estimate the overall effect along the socioeconomic gradient. Percentile bootstrapping was used to estimate confidence intervals and the proportion of the total effect mediated. Following Efron & Tibshirani’s (1993) methodology [40], data were resampled with replacement to generate an empirical distribution of mediation effects. The 95% confidence interval was then calculated using the 2.5^th^ and 97.5^th^ percentiles of this bootstrap distribution.

**Fig 1:**
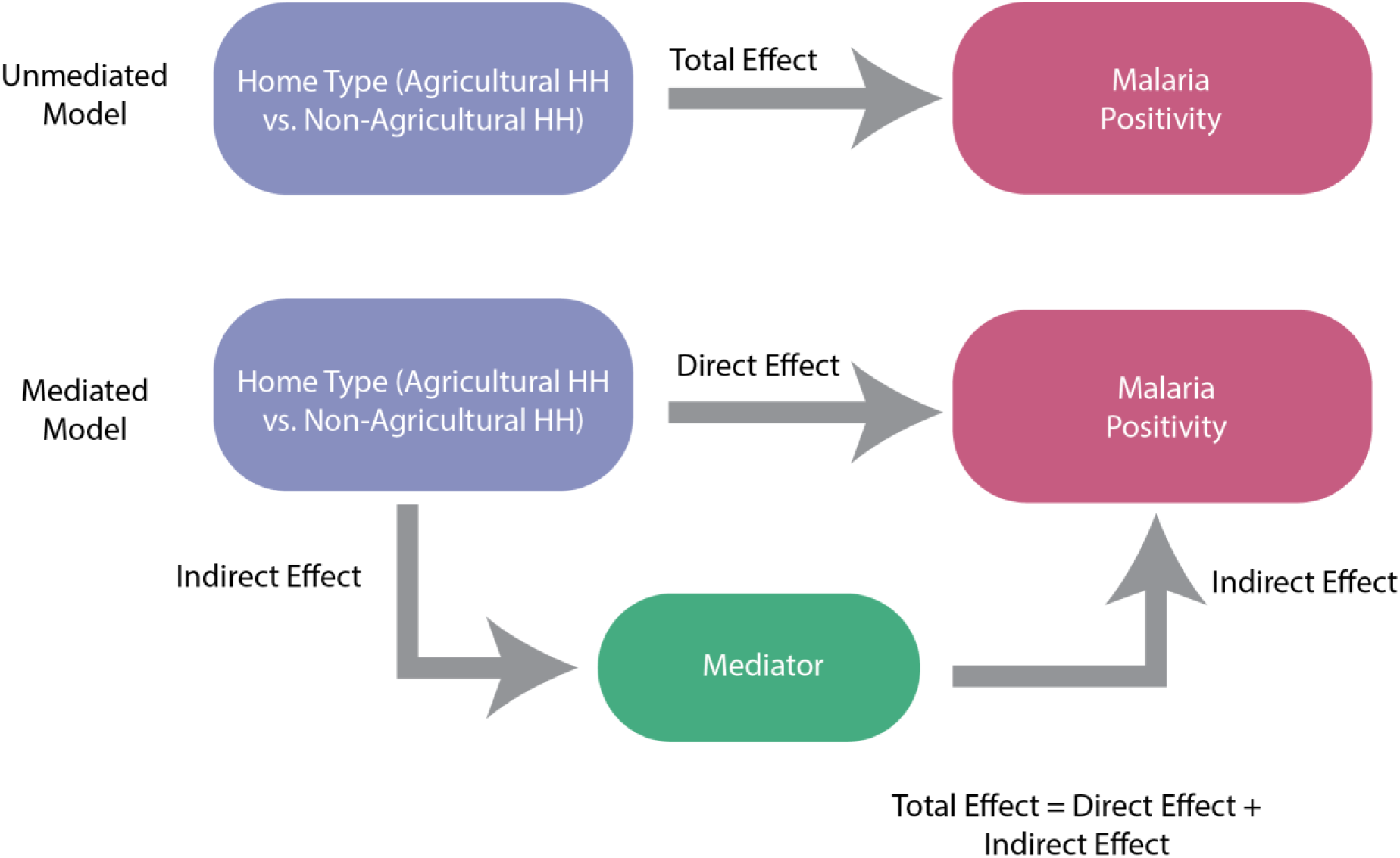
Mediation analysis model.

## Results

### Sample Overview and Descriptive Statistics

#### Sample Overview

Figure 2a depicts the country boundaries for the 15 countries included in this analysis, overlaid on a continental map of Africa. Nine of the countries are in Western Africa, four in Eastern Africa and two in Central Africa. Figure 2b provides a summary of the number of children tested for malaria using RDT or microscopy, alongside the proportion of rural residents in each country’s dataset in the most recent and preceding surveys, where available. Detailed counts are available in Supplementary Table 1. The 2018 Nigeria DHS tested the largest number of children under the age of five years (U5) for malaria using microscopy or RDT, with a total of 11,073 children. In contrast, the 2014 Ghana DHS had the smallest sample size, testing only 2,428 children (Supplementary Table 1a–b). In all countries studied, a higher proportion of data was collected from rural areas, except for Angola, where only 41% of the data came from rural areas (Figure 2b).

**Fig. 2.**
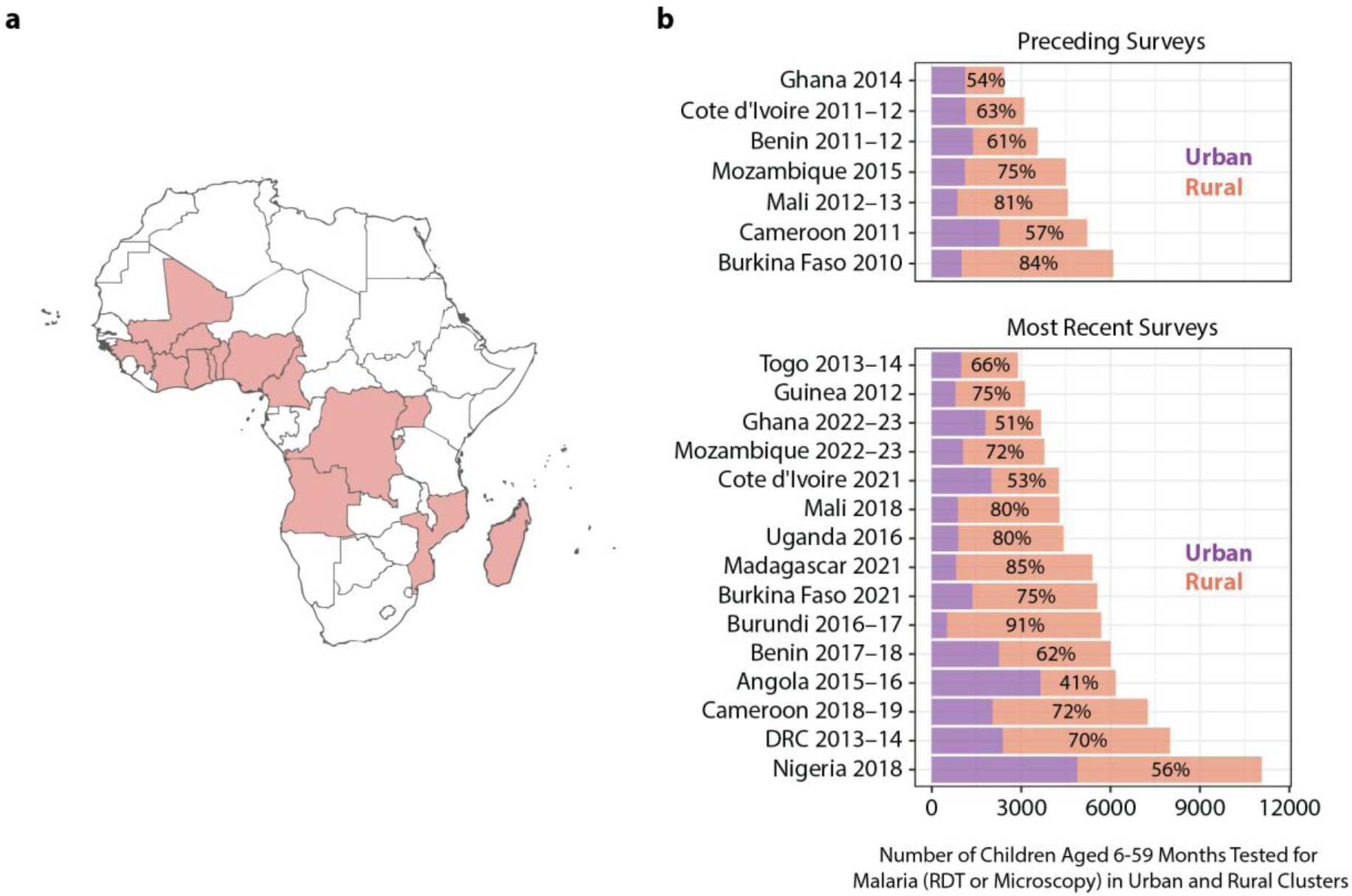
Countries included in the analysis, their location, survey years, and the number of U5 children tested in urban and rural areas. **(a)** Map of Africa highlighting the 15 countries whose DHS was included in this analysis. (**b)** Number of U5 children tested for malaria by RDT or microscopy, aggregated across urban (purple bars) and rural (orange bars) clusters from the most recent and preceding DHS where malaria and occupation data was collected. Fifteen countries had at least one survey and 7 had two surveys with the required data. The percentages shown represent rural clusters. Note: DRC refers to the Democratic Republic of the Congo.

#### Malaria Positivity and Net Use Variations in Agricultural Households and Non-Agricultural Worker Households

Malaria positivity rates were consistently higher in households where at least one adult was identified as an agricultural worker, overall and across urban and rural residences and countries (Supplementary Figure 1, Figure 3a–b, Supplementary Figure 2a–b). Overall, 30% of U5 children from agricultural worker households tested positive for malaria by RDT or microscopy, compared to 18% in non-agricultural households (Supplementary Figure 1). This disparity was particularly pronounced in urban areas, where malaria positivity rates were 2.5 times higher in agricultural households compared to non-agricultural households (25% vs. 10%, Figure 3a). Rural residents exhibited higher overall positivity rates for both home types, with a smaller difference between agricultural and non-agricultural households (31% vs. 26%, Figure 3b). Notably, this higher malaria prevalence in urban agricultural households was observed despite higher levels of bed net use among agricultural households (Figure 3c).

**Fig. 3.**
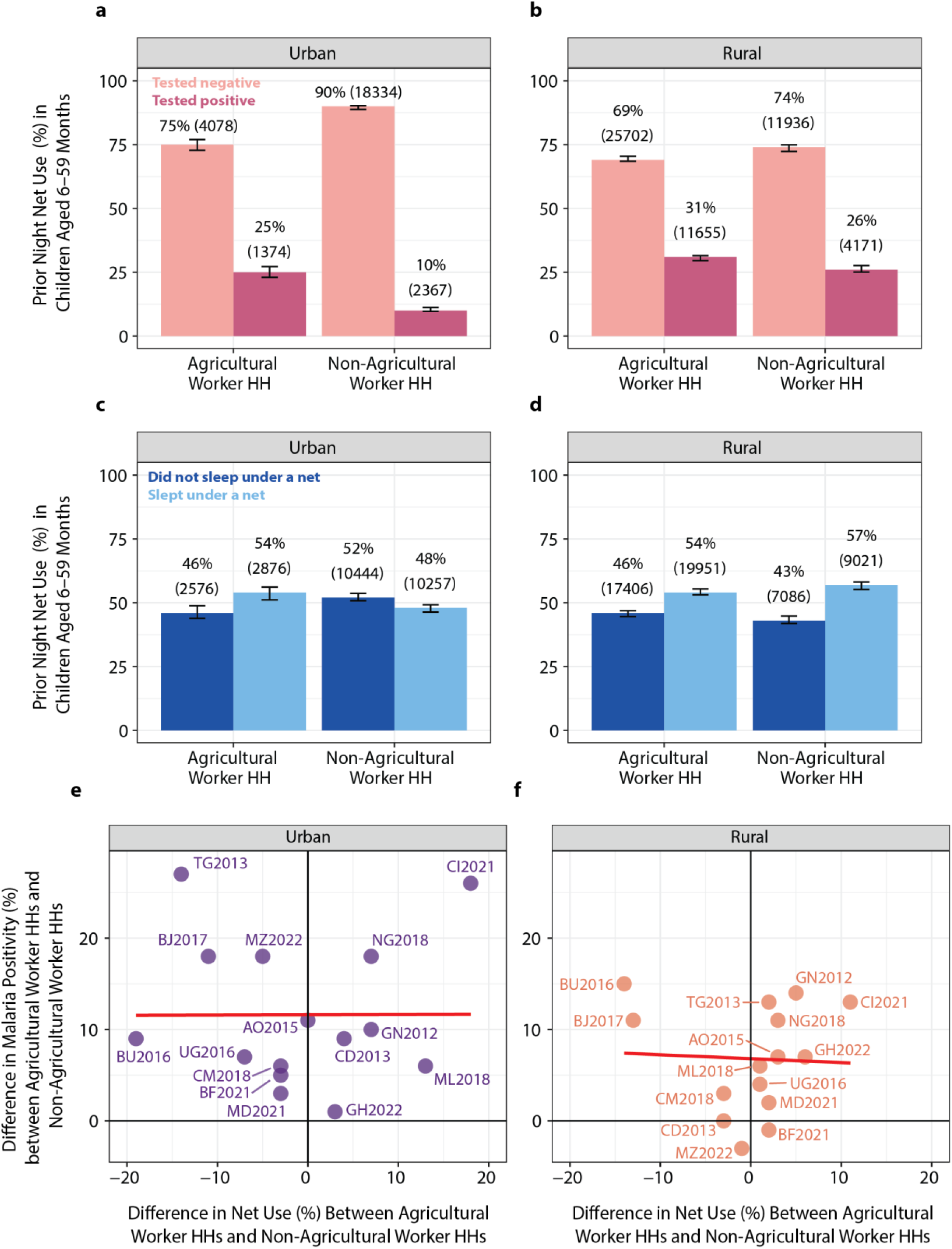
Malaria test results and net use among U5, by home type and residence type aggregated across the most recent DHS surveys in our dataset for all 15 countries. **(a)** Test positivity and negativity rates among children under five in households (HH) with agricultural and non-agricultural workers in urban areas. **(b)** Test positivity and negativity rates for the same categories in rural areas. **(c)** Net use rates among children under five in households with agricultural and non-agricultural workers in urban areas. **(d)** Net use rates among children under five in households with agricultural and non-agricultural workers in rural areas. Error bars represent 95% confidence intervals**. (e) and (f)** Country-specific differences in U5 malaria positivity rates and net use between agricultural worker households and non-agricultural worker households in urban (e) and (f) rural. The red line represents the linear trend between differences in net use and differences in malaria positivity. A positive difference implies that agricultural worker households have a greater malaria or net use rate relative to non-agricultural worker households **(a)** Urban, **(b)** Rural. **Country Abbreviations Key: AO** – Angola | **BF** – Burkina Faso | **BJ** – Benin | **BU** – Burundi | **CD** – Democratic Republic of the Congo | **CI** – Côte d’Ivoire | **CM** – Cameroon | **GH** – Ghana | **GN** – Guinea | **MD** – Madagascar **ML** – Mali | **MZ** – Mozambique | **NG – Nigeria** | **TG** – Togo | **UG** – Uganda.

At the country level, the highest urban malaria positivity rates were recorded in the 2017–18 Benin survey (47% in agricultural, 27% in non-agricultural households), while rural rates peaked in the 2012 Guinea survey (55% in agricultural, 40% in non-agricultural households) (Supplementary Figure 2a–b). Urban net use was highest in the 2017–18 Benin survey (71% in agricultural, 84% in non-agricultural) while in rural areas, net use was highest in the Mali 2018 survey (80% in agricultural and 79% in non-agricultural households). The lowest net use in both urban and rural areas were reported in the Angola 2015 – 16 survey (Supplementary Figure 3a–b). Country-level differences in malaria test positivity rate and net use between agricultural and non-agricultural worker households are shown in Figure 3e–f. In urban areas, the most pronounced disparities in malaria positivity between agricultural and non-agricultural households were observed in the 2013–14 Togo (27%-point difference) and 2021 Côte d’Ivoire (26%-point difference) surveys. Notably, higher net usage among urban children from non-agricultural worker households compared to those from agricultural worker households was noted in Togo (−14%-point difference, refer to Figure 3e). Conversely, the 2021 Côte d’Ivoire survey indicated greater bed net use among urban children in agricultural worker households than their counterparts in non-agricultural worker households (18%-point difference). In rural areas, disparities in test positivity rates between the two home types was greatest in the 2016 Burundi survey with a higher test positivity rate in agricultural worker households (15%-point difference, refer to Figure 3f) amid lower net use rates among agricultural households (−14%-point difference) and in the Guinea 2012 survey with 14%-point positive difference in test positivity rates amid greater net use (5%-point difference) in agricultural worker relative to non-agricultural worker households. The relationship between differences in net use and malaria positivity in countries was mostly flat in both urban and rural areas (red line in Figure 3e and f)

#### Heterogeneous Trends in Malaria Positivity vs. Net Use Rate Across Countries

To understand how malaria positivity varies with net use trends among children under five in agricultural and non-agricultural worker households, data from countries with at least two surveys and the required data were visualized (Figure 4). Burkina Faso and Cameroon exhibited a consistent pattern of decreasing malaria positivity rates alongside increasing net use rates over time, regardless of whether households were urban or rural, or agricultural or non-agricultural. In urban Burkina Faso and Cameroon, changes in the point estimate for net use, in the two studied groups, in the preceding and most recent survey were roughly similar. However, malaria positivity rate was higher in agricultural worker households in the Burkina Faso and Cameroon’s preceding survey, but these values trended downwards, converging to roughly the same estimate as non-agricultural worker households.

**Fig. 4:**
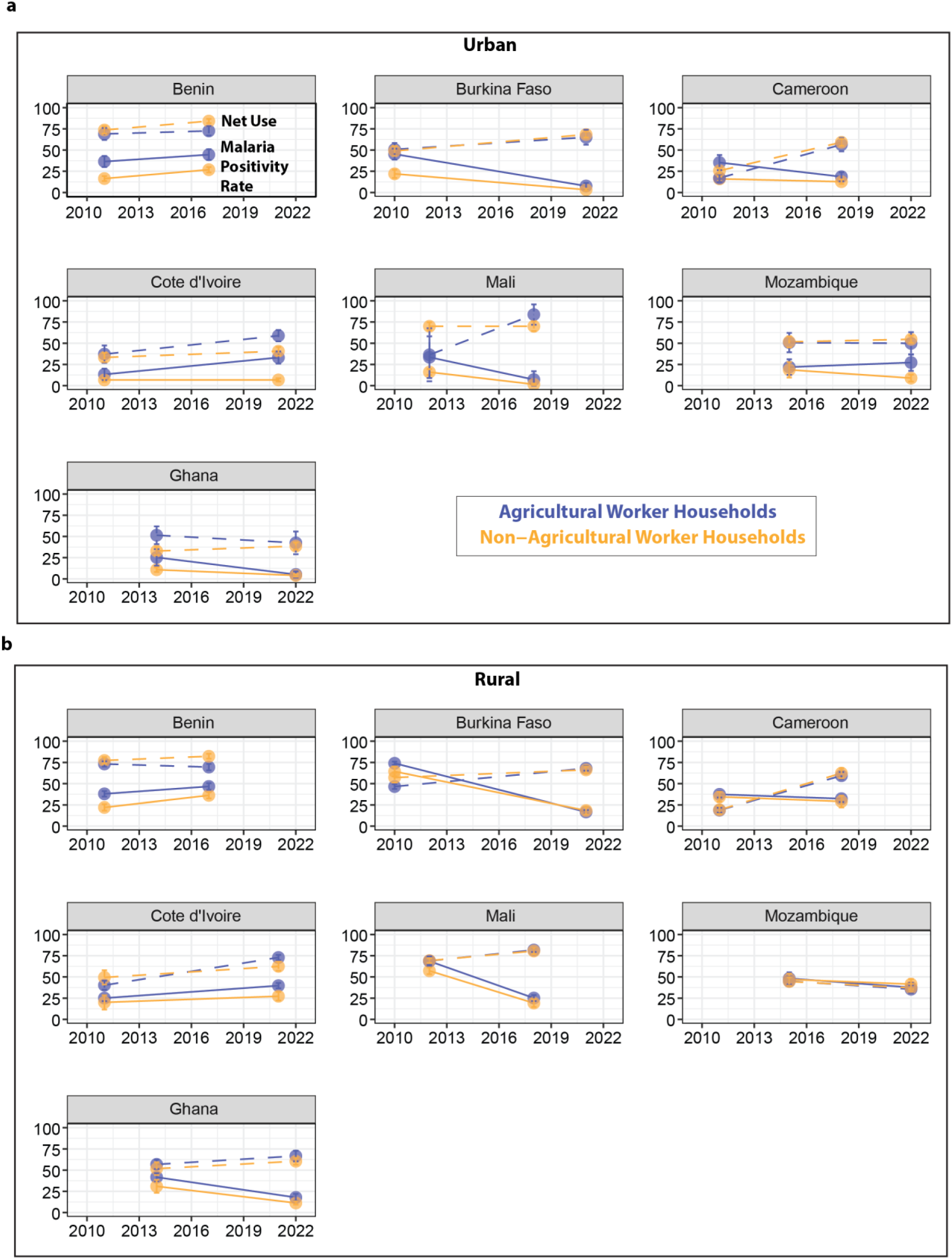
Change in malaria test positivity rate and net use between preceding survey and most recent survey among U5. **(a)** Rates in urban areas. **(b)** Rates in rural areas. Dashed lines represent net use rates and solid lines represent malaria test positivity rate in children under five. Error bars represent 95% confidence intervals.

Mali showed a similar pattern of increasing net use and decreasing malaria test positivity over time in rural areas among both agricultural and non-agricultural worker households. In urban Mali, net use remained relatively high and stable over time in non-agricultural worker households and trended upward in agricultural households, while malaria positivity rates declined. Urban Benin and rural Côte d’Ivoire showed an increasing malaria positivity rate despite rising net use rates for both home types. This unexpected trend raises questions about other factors influencing malaria transmission in these regions. In rural Benin, a patten of increasing malaria positivity coincided with a notable decrease of 3.7 percentage points in net use rates among agricultural households between the two survey years. In urban Côte d’Ivoire, malaria prevalence remained stable in non-agricultural households but rose agricultural households, although net use increased.

Both home types in urban Mozambique demonstrated relatively stable and moderate net use rates between the two survey years while malaria positivity declined in non-agricultural worker households but not in agricultural worker households (5.3 percentage point increase). However, rural Mozambique experienced a decline in both malaria positivity rates and net use rates for agricultural and non-agricultural households. In Ghana, urban agricultural households saw declines in both net use (9.0 percentage points) and malaria positivity (20.1 percentage points). Among Ghanaian urban non-agricultural households, net use increased slightly (5.7 percentage points) and malaria positivity declined slightly (6.7 percentage points). In rural areas in Ghana, net use increased (9.9 percentage points in agricultural households and 8.8 percentage points in non-agricultural households), while malaria positivity decreased in both groups (23.9 and 19.7 percentage points, respectively).

Supplementary Figure 4 shows the percentage change in bed nets distributed each year compared to the first recorded year, which provides additional context for interpreting malaria trends. In Benin and Côte d’Ivoire, positivity remains high despite consistent or increased net distribution.

#### Geographic Distribution of Urban Agricultural Worker Households

Given the remarkably higher under-five malaria prevalence in agricultural worker households compared to non-agricultural worker households in urban areas, we illustrate the geographic distribution of agricultural households in urban clusters—both individually and collectively— across the 15 countries included in our analysis to provide insight into administrative divisions where targeted interventions for this risk group may be needed (Figure 5). The maps reveal that the most populous administrative subdivisions tend to have a lower proportion of agricultural households in urban areas. However, this trend is not universal. In several countries, including Burundi, Democratic Republic of Congo (DRC), Côte d’Ivoire and Madagascar, even some of the most populous administrative subdivisions maintain a substantial urban agricultural worker presence, with 40–60% of households engaged in agricultural work.

**Fig. 5.**
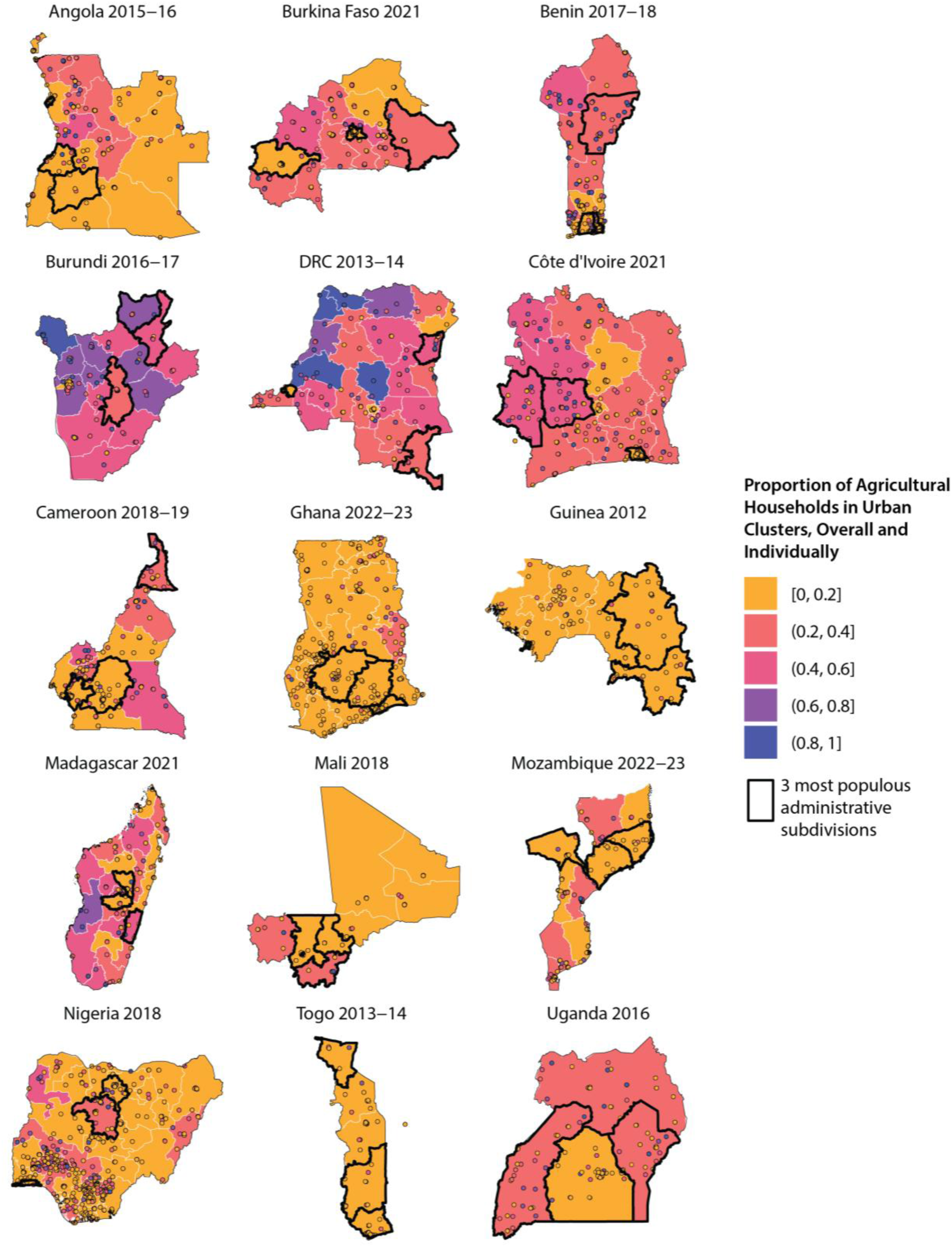
Distribution of urban agricultural worker households across geographic subdivisions. Maps show the proportion of agricultural worker households at the subdivision level (e.g. state, region), with the three most populous first-level administrative subdivisions in each country outlined in black. Points represent the proportion of agricultural households in each survey cluster. Only urban data included. First-level subdivisions in most countries are regions, except for these: Angola, Burundi, DRC, and Mozambique: provinces; Côte d’Ivoire: districts; Nigeria: states; Benin: departments. See Supplementary Table 6 for raster-derived population estimates for the three most populous first-level administrative subdivisions in each country.

Individual, household and environmental characteristics of children in agricultural and non-agricultural households.

Figure 6 illustrates the distribution of characteristics of children residing in agricultural and non-agricultural worker households in urban and rural settings from the most recent DHS surveys in the 15 countries examined. Compared to non-agricultural worker households and irrespective of place of residence, agricultural worker households exhibited larger mean household sizes (7.88 members vs. 6.57 in urban, 7.52 vs. 6.55 in rural), and resided in communities with higher enhanced vegetation indices (EVI) (0.373 vs. 0.288 in urban, 0.379 vs. 0.354 in rural), and higher prevalence of poor housing quality (58% vs. 26% in urban, 84% vs. 62% in rural), poverty (19% vs. 2% in poorest wealth quintile in urban, 34% vs. 22% in poorest wealth quintile in rural), child stunting (13.2% vs. 8.0% in urban, 18.9% vs. 13.8% in rural), and higher prevalence of children living in clusters with low treatment-seeking for fever (62% vs. 49% in urban areas, 63% vs. 50% in rural areas). Agricultural worker households also resided in locations with greater precipitation levels in urban but not in rural areas (155 mm/month vs. 147 in urban). Distributions of relative humidity, temperature, gender, and age were similar between all groups. Urban-rural comparisons of agricultural worker households revealed minimal differences in household size, EVI, precipitation, relative humidity, temperature, age, and gender distributions. However, both agricultural and non-agricultural workers in rural areas demonstrated higher prevalence of poor housing quality, poverty, and stunting compared to their urban counterparts.

**Fig. 6.**
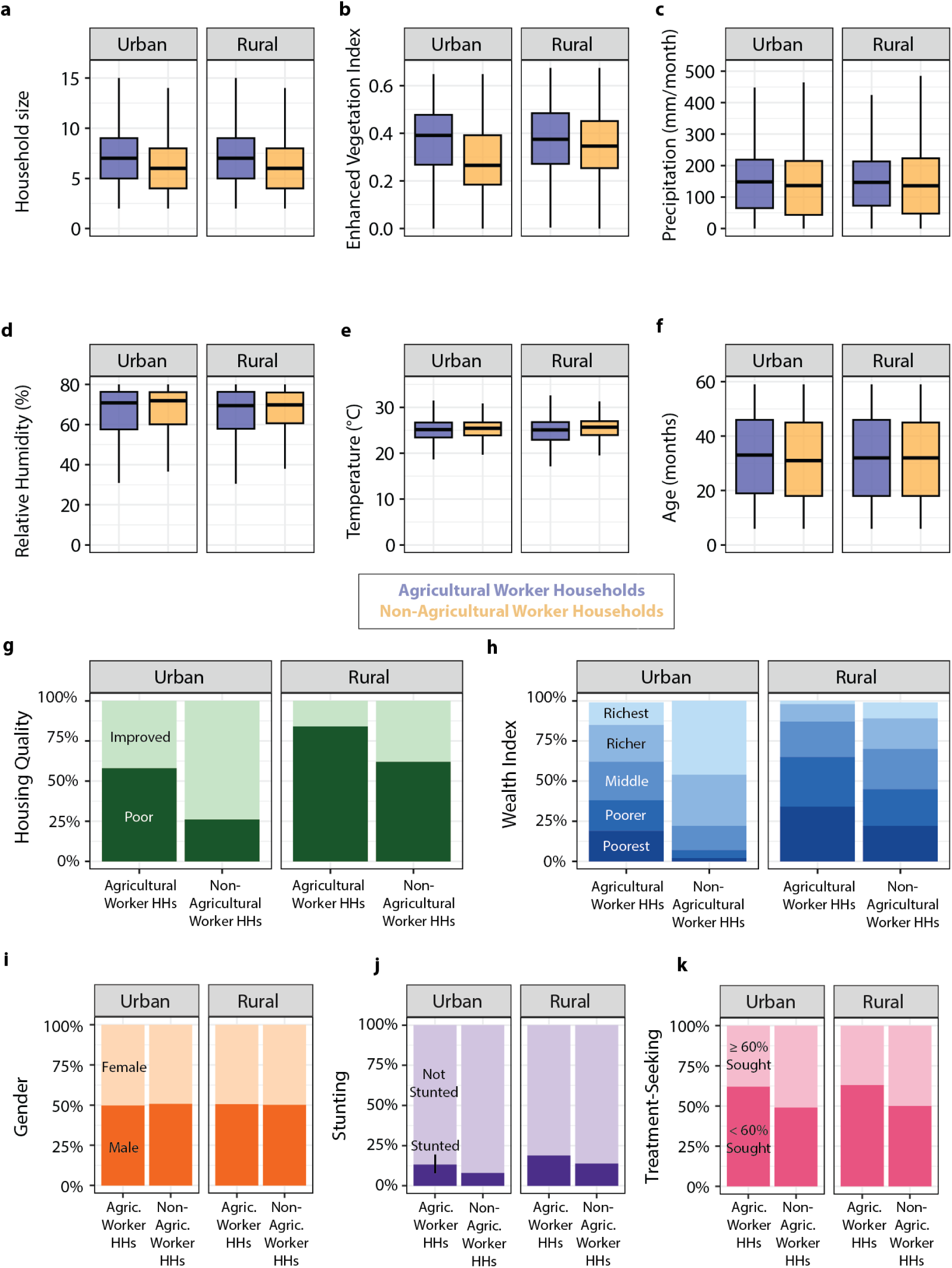
Characteristics of the study population in the 15 most recent DHS surveys in our dataset. (**a–f)** Boxplots showing the distribution of continuous and discrete covariates **(a)** household size, **(b)** enhanced vegetation index, **(c)** monthly precipitation, **(d)** percent relative humidity, **(e)** temperature in °C, and **(e)** age in months, separated by agricultural worker households (HH) (blue) and non-agricultural worker households (yellow). **g–k.** Percentage distribution of categorical covariates **(g**) Improved and poor housing quality, **(h)** wealth quintiles, **(i)** gender, **(j)** stunting, and **(k)** treatment-seeking for fever, classified as “≥60% sought treatment” if either no one reported fever (assumed high treatment-seeking) or at least 60% of those with fever sought care; otherwise, they are classified as “<60% sought treatment.”

Impact of potential mediators on the relationship between home type (agricultural vs. non-agricultural) and malaria positivity as measured by percent mediation.

In all urban settings of the 15 countries studied, wealth emerged as the strongest mediator, accounting for 46.45% (95% CI: 41.74%–51.78%) of the relationship between home type and malaria positivity. EVI was the second most influential mediator, explaining 27.41% (95% CI: 24.38%–30.70%) of the relationship. Housing quality and treatment-seeking also played influential roles, mediating 14.06% (95% CI: 11.79%–16.52%) and 13.99% (95% CI: 11.65%–16.51%), respectively, of the relationship. Other significant mediators included household size (3.19%, 95% CI: 1.99%–4.50%) and stunting (2.70%, 95% CI: 1.88%–3.57%). The mediating effect of net use (−0.09%, 95% CI: −0.33%–0.10%) was not statistically significant (Figure 7).

**Fig. 7:**
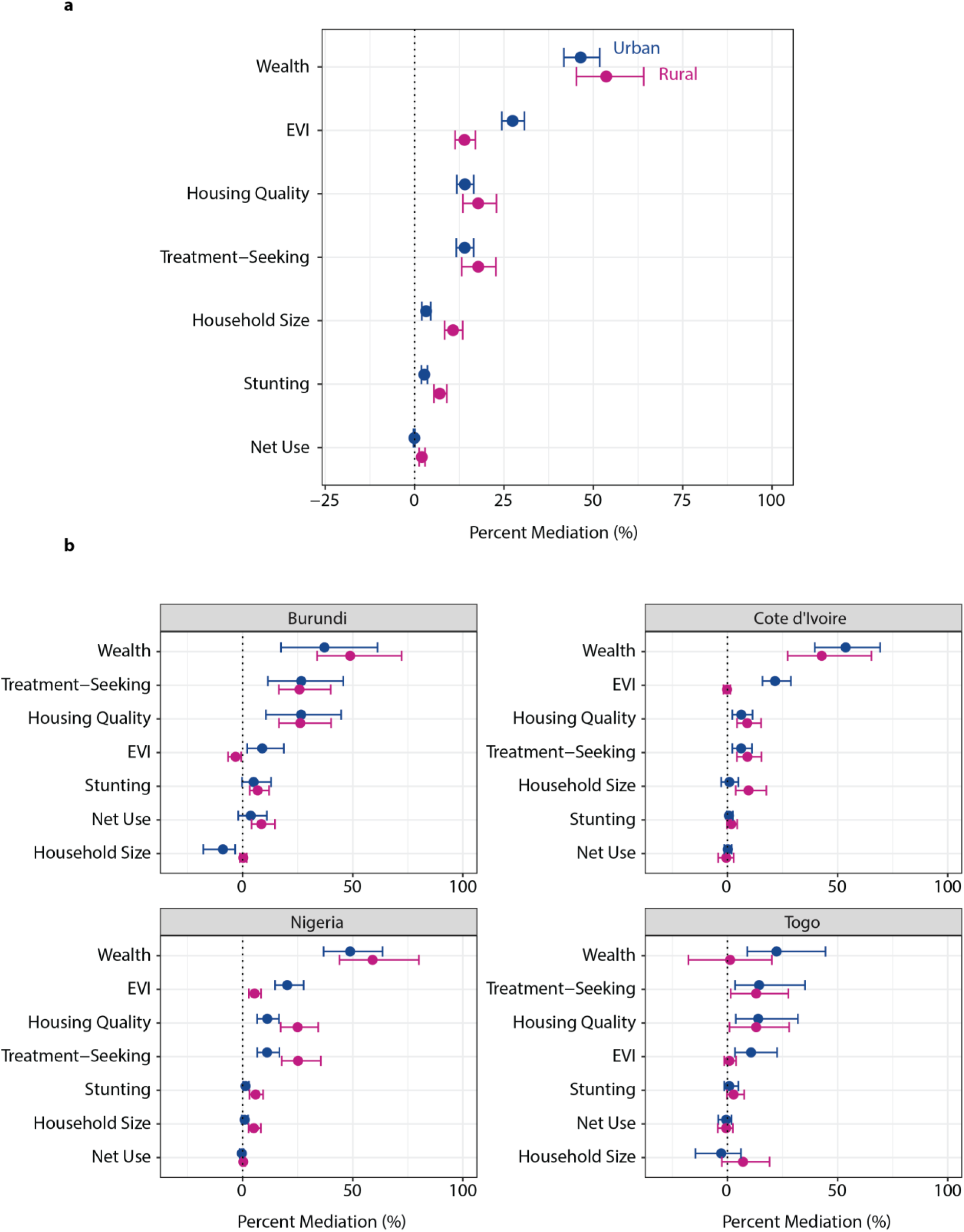
Mediators’ contributions to the home type-malaria positivity relationship, measured by percent mediation in (**a)** all 15 countries using their most recent survey in our dataset **and (b)** countries with stable and interpretable estimates. Percent mediation and confidence intervals are calculated using bootstrapping. Error bars represent 95% confidence intervals.

In rural settings, wealth remained the most substantial mediator, explaining 53.61% (95% CI: 45.25%–64.12%) of the relationship between home type and malaria positivity. Treatment-seeking played a stronger mediating role than in urban areas, explaining 17.78% (95% CI: 13.16%–22.73%) of the relationship. Housing quality accounted for 17.76% (95% CI: 13.49%–22.89%) of the relationship. Compared to urban areas, EVI extracted for rural areas exhibited about half the strength of the mediating effect (13.93%, 95% CI: 11.30%–16.99%). Household size played a more significant role in rural areas, mediating 10.74% (95% CI: 8.37%–13.44%) of the relationship. Stunting was another significant mediator (7.01%, 95% CI: 5.39%–8.98%). Net use had a small mediating effect (2.00%, 95% CI: 1.26%–2.89%) (Figure 7).

In both urban and rural settings, direct effects remained statistically significant, indicating that being in an agricultural household is associated with higher odds of testing positive for malaria through pathways not mediated by the variable under evaluation. This suggests additional mechanisms independent of the examined mediator (Supplementary Table 4).

Country-level variations in the percentage mediated by various factors are presented in Figure 7b. While results were generated for all 15 countries included in the study (Supplementary figure 5), we observed that the confidence intervals for most of the countries were very wide, indicating substantial uncertainty in these estimates. To ensure clarity and interpretability, we present forest plots of percent mediation for the four countries with more stable and interpretable estimates (that is, narrower confidence intervals).

The four countries highlighted (Burundi, Côte d’Ivoire, Nigeria, and Togo), exhibited similar patterns in percent mediation, with wealth, EVI, housing quality, and treatment-seeking generally emerging as the strongest mediators. Notably, wealth was a weaker mediator in Togo compared to the other three countries. For all four countries, the mediators stunting, net use, and household size had small percentage mediations, with estimates near zero and relatively narrow confidence intervals.

## Discussion

### Overview of findings and implications

As national malaria programs in endemic countries re-prioritize in response to shrinking funding for control and elimination, locally relevant information becomes increasingly vital, particularly data on who is at risk, where they live, their characteristics, and the uptake and effectiveness of malaria interventions. Additionally, programs are likely to increasingly adopt horizontal programming approaches that use resources more efficiently, given the shared risk factors for malaria and other diseases. This study aimed to generate evidence to inform such strategies.

Our analysis revealed significant disparities in childhood malaria prevalence between urban agricultural and non-agricultural households. These differences were especially pronounced in urban areas of Togo, Côte d’Ivoire, Benin, Mozambique, and Nigeria. We also observed variations in net use patterns: in Côte d’Ivoire, urban agricultural households reported higher rates of net use compared to their non-agricultural counterparts, whereas the opposite was true in Togo.

Paradoxically, in Côte d’Ivoire, malaria prevalence among children in urban agricultural households increased even as net use rose, while prevalence remained constant in non-agricultural households. Benin showed similar findings, although prevalence increased across all groups. This counterintuitive trend was clarified by our finding that net use does not explain the relationship between home type and malaria positivity in urban agricultural settings, possibly due to limited indoor night-time biting, insecticide resistance or higher infectious outdoor biting. This finding does not mean nets are not helpful but could imply the need for complementary interventions due to multiple interacting risk factors.

As indicated by the EVI, environmental conditions were found to be a leading driver of malaria burden for children in these areas. As such, malaria programs in countries where agricultural workers have a notable presence in urban areas should give serious consideration to identifying and addressing environmental determinants of malaria.

Interestingly, we observed marked differences in mediation estimates across urban and rural settings within certain countries. In Nigeria, housing quality and treatment-seeking behaviors accounted for a notably higher proportion of the mediated effect in urban areas compared to rural. This suggests that structural housing improvements and access to care may be particularly impactful in urban agricultural communities. In Côte d’Ivoire, household size emerged as a more substantial mediator in rural settings, indicating potential crowding-related transmission dynamics that warrant further investigation. These differences reinforce the importance of subnational tailoring of malaria control strategies based on local risk profiles.

Our study also highlights that households engaged in urban agriculture often experience high levels of poverty, putting them at risk of other endemic infectious diseases. Therefore, to maximize the impact of disease control efforts, integrated approaches that combine malaria prevention with environmental management, improved treatment-seeking behaviors, housing upgrades, poverty reduction, and nutritional support may be more effective. As national malaria control programs increasingly collaborate with other sectors to support subnational targeting [41], a next step would be the multisectoral co-creation of *Health for All Policies* strategies [42].

### Comparison to research literature

Although our study findings provide a unique overview of country-specific childhood malaria risk stratified by resident adult occupation in the agricultural and non-agricultural sectors and the mediators of this relationship, its findings complement and align with the research literature. In their 2022 analysis of DHS data combined with land cover and land use datasets, Shah and colleagues found that under-five children situated near forest cover, rainfed cropland in rural and irrigated or post-flooding in urban areas were more likely to test positive for malaria [16]. Beke’s work, on the other hand, among children whose parents engaged in agricultural activities from six sites equally split by urban and rural in Cote d’Ivoire provided a granular view showing that irrigated sites were important in rural malaria transmission and wetlands in urban areas [14]. This analysis adds to this body of work by highlighting the value of EVI, a measure of vegetation greenness, indicative of rainfall frequency, forest cover or farmland, in identifying agricultural communities at risk of malaria. In addition, it highlights ways in which agricultural communities differ from non-agricultural communities in terms of housing quality, stunting and treatment seeking, important mediators of malaria risk and possibly other diseases, which would enable health programs to design more comprehensive and integrated intervention strategies.

Other studies have found that the use of bed nets is not associated with malaria risk in urban areas [16,20,43]. This work sheds some light on this issue, showing that although children in urban agricultural households have an elevated malaria risk, bed nets do not mediate the effect of agricultural work on childhood malaria, even though children in those households, with some exceptions, use nets as the same or even greater levels than non-agricultural worker households. It is possible that the presence of members of agricultural households in study datasets obscures the impact of net use in urban areas.

This analysis also show that agricultural worker households are not different from other communities in terms of the factors highly correlated with increased malaria risk: Lower wealth Index [20,44,45], higher EVI [20,46], poorer housing quality [3,4], inadequate treatment seeking behaviors in febrile patients[47,48] and larger household size [27] have been found to explain malaria risk in studies conducted in the general population. The relationship between malaria risk and stunting is not well understood but appears to be bidirectional [49,50].

### Limitations

This study has several limitations related to its design, data availability and resolution, and timing of data collection. First, the reliance on cross-sectional data limits our ability to infer causality. While we examined single mediator pathways, it is possible that some mediators lie on the causal pathway of others, indicating more complex, sequential relationships that were not examined in this analysis. In addition, although we accounted for mediators such as household wealth and housing quality, we were unable to incorporate micro-level environmental factors—such as proximity to mosquito breeding sites or land use patterns— which likely vary across household types and may meaningfully influence malaria transmission.

We also lacked specific information on child-level exposures, such as whether children in agricultural households accompanied adults to worksites, as well as the extent to which these children spent time outdoors or near potential vector habitats, which could affect individual malaria risk. It was also unclear whether adults engaged in agricultural work near their homes or in more distant locations, adding further uncertainty. The 27km resolution of the temperature and relative humidity data may be unsuitable to capture differences between agricultural and non-agricultural worker communities. Furthermore, urban and rural classifications were based on administrative definitions, which may have misclassified some areas and, consequently, biased estimates of malaria risk. Finally, our analysis was limited to countries with both malaria and occupation data available during specific survey years, which may restrict the generalizability of our findings.

### Recommendations for future studies

Future malaria indicator surveys should capture information on employment in the agricultural sector, daily work times and seasonality to enable continued surveillance of malaria burden in this population. Studies examining children’s exposure to agriculture, such as time spent in fields with their parents, could provide further insight. Studies should also investigate what sorts of interventions would work best for residents of agricultural communities and how these would vary by age and urban and rural places of residence.

Longitudinal and modeling studies would be valuable to assess changes in malaria prevalence and risk factors over time, evaluate intervention impact, and identify emerging transmission trends. As climate change continues to reshape malaria dynamics, particularly in agricultural regions, advanced tools like remote sensing could play a pivotal role in identifying high risk areas and improving the targeting of interventions.

### Conclusions

This study reveals that children in agricultural households in urban settings face significantly higher malaria risks compared to those in non-agricultural households. It also finds that while vegetation indices mediate the associations between agricultural work and malaria risk, nets had limited impact. Given country-specific differences in risk factors, integrated and locally relevant interventions are crucial for reducing malaria vulnerability, particularly among vulnerable groups in both urban and rural agricultural settings.

## List of abbreviations

DHS - Demographic and Health Survey

EVI - Enhanced Vegetation Index

GADM - Database of Global Administrative Areas

IRB - Institutional Review Board

LLINs - Long-lasting Insecticidal Nets

RDT – Rapid Diagnostic Tests

U5 - children under the age of five years

## Declarations

### Ethics approval and consent to participate

This study used publicly available, de-identified data from the 2010–2023 DHS, which can be found at: https://dhsprogram.com/data/available-datasets.cfm. Ethical clearance for the DHS protocols was obtained from ICF International’s Institutional Review Board (IRB). The surveys ensured compliance with international ethical standards for informed consent, voluntary participation, privacy, and confidentiality. The datasets were requested, and a letter of data authorization was received from ICF International through the DHS Program. More details regarding ethical standards of the DHS data are available at: https://www.dhsprogram.com/methodology/Protecting-the-Privacy-of-DHS-Survey-Respondents.cfm

### Consent for publication

Not applicable

### Availability of data and materials

The datasets generated and analyzed during the current study are available through Zenodo via the urban-malaria/Agricultural-manuscript: Agriculture-manuscript v1.0 repository, https://doi.org/10.5281/zenodo.15831265

### Authors’ contributions

IDO conceptualized the study and its design, identified funding for the study, and led data curation, analysis, interpretation and writing. GL and CC supported data analysis, interpretation and writing. LM provided feedback on the study design and reviewed the outputs of the data analysis and drafts of the study writeup.

## Acknowledgements

We acknowledge members of the Urban Malaria Lab who provided feedback on the analysis, including Eniola Bamgboye, Hephzibah Adeniji, Yusuf Jamiu, and Gift Enang.

## References

1. World malaria report 2024: addressing inequity in the global malaria response [Internet]. Geneva: World Health Organization; Available from: https://www.who.int/teams/global-malaria-programme/reports/world-malaria-report-2024

2. Yang D, He Y, Wu B, Deng Y, Li M, Yang Q, et al. Drinking water and sanitation conditions are associated with the risk of malaria among children under five years old in sub-Saharan Africa: A logistic regression model analysis of national survey data. J Adv Res. 2020;21:1–13.

3. Tusting LS, Bottomley C, Gibson H, Kleinschmidt I, Tatem AJ, Lindsay SW, et al. Housing Improvements and Malaria Risk in Sub-Saharan Africa: A Multi-Country Analysis of Survey Data. Von Seidlein L, editor. PLoS Med. 2017;14:e1002234.

4. Tusting LS, Ippolito MM, Willey BA, Kleinschmidt I, Dorsey G, Gosling RD, et al. The evidence for improving housing to reduce malaria: a systematic review and meta-analysis. Malar J. 2015;14:209.

5. Bhatt S, Weiss DJ, Cameron E, Bisanzio D, Mappin B, Dalrymple U, et al. The effect of malaria control on Plasmodium falciparum in Africa between 2000 and 2015. Nature. 2015;526:207–11.

6. Malm KL, Peprah NY, Mohammed W, Adomako B-Y, Oppong S, Boateng P, et al. A retrospective analysis of malaria deaths in the pre- and intra-COVID 19 pandemic era, Ghana, 2016–2021. Ampofo GD, editor. PLoS ONE. 2024;19:e0286212.

7. Heuschen A-K, Lu G, Razum O, Abdul-Mumin A, Sankoh O, Von Seidlein L, et al. Public health-relevant consequences of the COVID-19 pandemic on malaria in sub-Saharan Africa: a scoping review. Malar J. 2021;20:339.

8. Kuppalli K, Dunning J, Damon I, Mukadi-Bamuleka D, Mbala P, Ogoina D. The worsening mpox outbreak in Africa: a call to action. The Lancet Infectious Diseases. 2024;24:1190–2.

9. Rodríguez-Morales AJ, Luna C, Flores-Girón L, Membrillo de Novales FJ, Torres-Martinez C, Camacho-Moreno G, et al. Mpox in children (2024): New Challenges. BMJ Paediatr Open. 2024;8:e003030.

10. Yu Q, Qu Y, Zhang L, Yao X, Yang J, Chen S, et al. Spatial spillovers of violent conflict amplify the impacts of climate variability on malaria risk in sub-Saharan Africa. Proc Natl Acad Sci USA. 2024;121:e2309087121.

11. Coalson JE, Anderson EJ, Santos EM, Madera Garcia V, Romine JK, Luzingu JK, et al. The Complex Epidemiological Relationship between Flooding Events and Human Outbreaks of Mosquito-Borne Diseases: A Scoping Review. Environ Health Perspect. 2021;129:096002.

12. Umar N, Gray A. Flooding in Nigeria: a review of its occurrence and impacts and approaches to modelling flood data. International Journal of Environmental Studies. 2023;80:540–61.

13. Symons TL, Lubinda J, McPhail M, Saddler A, Van Den Berg M, Baggen H, et al. Estimating the potential malaria morbidity and mortality avertable by the President’s Malaria Initiative in 2025: a geospatial modelling analysis [Internet]. Infectious Diseases (except HIV/AIDS); 2025 [cited 2025 Jun 19]. Available from: http://medrxiv.org/lookup/doi/10.1101/2025.02.28.25323072

14. Beke OA-H, Assi S-B, Kokrasset APH, Dibo KJD, Tanoh MA, Danho M, et al. Implication of agricultural practices in the micro-geographic heterogeneity of malaria transmission in Bouna, Côte d’Ivoire. Malaria Journal. 2023;22:313.

15. Chan K, Tusting LS, Bottomley C, Saito K, Djouaka R, Lines J. Malaria transmission and prevalence in rice-growing versus non-rice-growing villages in Africa: a systematic review and meta-analysis. The Lancet Planetary Health. 2022;6:e257–69.

16. Shah HA, Carrasco LR, Hamlet A, Murray KA. Exploring agricultural land-use and childhood malaria associations in sub-Saharan Africa. Sci Rep. 2022;12:4124.

17. Keiser J, De Castro MC, Maltese MF, Bos R, Tanner M, Singer BH, et al. Effect of irrigation and large dams on the burden of malaria on a global and regional scale. Am J Trop Med Hyg. 2005;72:392–406.

18. Fornace KM, Diaz AV, Lines J, Drakeley CJ. Achieving global malaria eradication in changing landscapes. Malar J. 2021;20:69.

19. Tepa A, Kengne-Ouafo JA, Djova VS, Tchouakui M, Mugenzi LMJ, Djouaka R, et al. Molecular Drivers of Multiple and Elevated Resistance to Insecticides in a Population of the Malaria Vector Anopheles gambiae in Agriculture Hotspot of West Cameroon. Genes. 2022;13:1206.

20. Leonard C, Ozodiegwu ID, Stahlfeld A, Chiziba C, Adam NS, Diallo OO, et al. Efficiencies in the allocation of insecticide-treated nets for malaria prevention in urban Sub-Saharan Africa [Internet]. Epidemiology; 2025 [cited 2025 Jun 19]. Available from: 10.1101/2025.05.29.25328568

21. The DHS Program - Demographic and Health Survey (DHS) [Internet]. [cited 2024 Oct 25]. Available from: https://dhsprogram.com/methodology/survey-Types/dHs.cfm

22. Burgert C, Colston J, Roy T, Zachary B. Geographic Displacement Procedure and Georeferenced Data Release Policy for the Demographic and Health Surveys. DHS Spatial Analysis Reports No. 7. Calverton, Maryland: ICF International; 2013.

23. Patz JA, Olson SH. Malaria risk and temperature: Influences from global climate change and local land use practices. Proc Natl Acad Sci USA. 2006;103:5635–6.

24. Ebhuoma O, Gebreslasie M. Remote Sensing-Driven Climatic/Environmental Variables for Modelling Malaria Transmission in Sub-Saharan Africa. IJERPH. 2016;13:584.

25. Ricotta EE, Frese SA, Choobwe C, Louis TA, Shiff CJ. Evaluating local vegetation cover as a risk factor for malaria transmission: a new analytical approach using ImageJ. Malar J. 2014;13:94.

26. Isiko I, Nyegenye S, Bett DK, Asingwire JM, Okoro LN, Emeribe NA, et al. Factors associated with the risk of malaria among children: analysis of 2021 Nigeria Malaria Indicator Survey. Malaria Journal. 2024;23:109.

27. Huldén L, McKitrick R, Huldén L. Average Household Size and the Eradication of Malaria. Journal of the Royal Statistical Society Series A: Statistics in Society. 2014;177:725–42.

28. Okiring J, Epstein A, Namuganga JF, Kamya EV, Nabende I, Nassali M, et al. Gender difference in the incidence of malaria diagnosed at public health facilities in Uganda. Malaria Journal. 2022;21:22.

29. Quaresima V, Agbenyega T, Oppong B, Awunyo JADA, Adomah PA, Enty E, et al. Are Malaria Risk Factors Based on Gender? A Mixed-Methods Survey in an Urban Setting in Ghana. Tropical Medicine and Infectious Disease. 2021;6:161.

30. Atusingwize E, Deane K, Musoke D. Social determinants of malaria in low- and middle-income countries: a mixed-methods systematic review. Malar J. 2025;24:165.

31. Worrall E, Basu S, Hanson K. Is malaria a disease of poverty? A review of the literature. Tropical Med Int Health. 2005;10:1047–59.

32. GADM [Internet]. [cited 2024 Nov 8]. Available from: https://gadm.org/

33. Uganda - Subnational Administrative Boundaries - Humanitarian Data Exchange [Internet]. [cited 2024 Nov 18]. Available from: https://data.humdata.org/dataset/cod-ab-uga?

34. WorldPop:: Population Counts [Internet]. [cited 2025 Mar 26]. Available from: https://hub.worldpop.org/geodata/listing?id=78

35. Huete A, Didan K, Miura T, Rodriguez EP, Gao X, Ferreira LG. Overview of the radiometric and biophysical performance of the MODIS vegetation indices. Remote Sensing of Environment. 2002;83:195–213.

36. Weiss DJ, Atkinson PM, Bhatt S, Mappin B, Hay SI, Gething PW. An effective approach for gap-filling continental scale remotely sensed time-series. ISPRS Journal of Photogrammetry and Remote Sensing. 2014;98:106–18.

37. Data Downloads CHIRPS 2.0 | Early Warning and Environmental Monitoring Program [Internet]. [cited 2025 Mar 12]. Available from: https://earlywarning.usgs.gov/fews/datadownloads/Global/CHIRPS%202.0

38. ERA5 monthly averaged data on pressure levels from 1940 to present [Internet]. [cited 2025 Mar 12]. Available from: https://cds.climate.copernicus.eu/datasets/reanalysis-era5-pressure-levels-monthly-means?tab=overview

39. World malaria report 2023 [Internet]. [cited 2024 Oct 18]. Available from: https://www.who.int/publications/i/item/9789240086173

40. Efron B, Tibshirani R. An introduction to the bootstrap. Nachdr. Boca Raton, Fla.: Chapman & Hall; 1998.

41. Onyango L, Ouédraogo-Ametchie G, Ozodiegwu I, Galatas B, Gerardin J. Subnational tailoring of malaria interventions for strategic planning and prioritization: Experience and perspectives of five malaria programs. Ashton R, editor. PLOS Glob Public Health. 2025;5:e0003811.

42. Greer SL, Falkenbach M, Siciliani L, McKee M, Wismar M, Figueras J. From Health in All Policies to Health for All Policies. The Lancet Public Health. 2022;7:e718–20.

43. Mathanga DP, Tembo AK, Mzilahowa T, Bauleni A, Mtimaukenena K, Taylor TE, et al. Patterns and determinants of malaria risk in urban and peri-urban areas of Blantyre, Malawi. Malaria Journal. 2016;15:590.

44. Sarfo JO, Amoadu M, Kordorwu PY, Adams AK, Gyan TB, Osman A-G, et al. Malaria amongst children under five in sub-Saharan Africa: a scoping review of prevalence, risk factors and preventive interventions. Eur J Med Res. 2023;28:80.

45. Anjorin S, Okolie E, Yaya S. Malaria profile and socioeconomic predictors among under-five children: an analysis of 11 sub-Saharan African countries. Malar J. 2023;22:55.

46. Okunlola O, Oloja S, Ebiwonjumi A, Oyeyemi O. Vegetation index and livestock practices as predictors of malaria transmission in Nigeria. Sci Rep. 2024;14:9565.

47. Omondi CJ, Odongo D, Otambo WO, Ochwedo KO, Otieno A, Lee M-C, et al. Malaria diagnosis in rural healthcare facilities and treatment-seeking behavior in malaria endemic settings in western Kenya. Zwerling A, editor. PLOS Glob Public Health. 2023;3:e0001532.

48. Workineh B, Mekonnen FA. Early treatment-seeking behaviour for malaria in febrile patients in northwest Ethiopia. Malar J. 2018;17:406.

49. Gebreegziabher E, Dah C, Coulibaly B, Sie A, Bountogo M, Ouattara M, et al. The Association between Malnutrition and Malaria Infection in Children under 5 Years in Burkina Faso: A Longitudinal Study. The American Journal of Tropical Medicine and Hygiene. 2023;108:561–8.

50. Das D, Grais RF, Okiro EA, Stepniewska K, Mansoor R, Van Der Kam S, et al. Complex interactions between malaria and malnutrition: a systematic literature review. BMC Med. 2018;16:186.

