## Supplementary files for "Agricultural Work, Malaria Prevalence, and Mediating Factors: A Cross-Sectional Analysis of Data from 15 Sub-Saharan African Countries to Inform Risk Stratification and Intervention Planning"

**Supplementary Table 1.** Number of children aged 6–59 months tested for malaria by country and by place of residence.

**(a)** Preceding Surveys

| **DHS** | **Urban** | **Rural** | **Total** |
| --- | --- | --- | --- |
| Burkina Faso 2010 | 5096 | 994 | 6090 |
| Cameroon 2011 | 2947 | 2268 | 5215 |
| Mali 2012–13 | 3714 | 852 | 4566 |
| Mozambique 2015 | 3396 | 1109 | 4505 |
| Benin 2011–12 | 2175 | 1379 | 3554 |
| Cote d'Ivoire 2011–12 | 1970 | 1135 | 3105 |
| Ghana 2014 | 1310 | 1118 | 2428 |

### **(b)** Recent Surveys

| **DHS** | **Urban** | **Rural** | **Total** |
| --- | --- | --- | --- |
| Nigeria 2018 | 6192 | 4881 | 11073 |
| DRC 2013–14 | 5616 | 2379 | 7995 |
| Cameroon 2018–19 | 5218 | 2034 | 7252 |
| Angola 2015–16 | 2530 | 3643 | 6173 |
| Benin 2017–18 | 3745 | 2254 | 5999 |
| Burundi 2016–17 | 5176 | 509 | 5685 |
| Burkina Faso 2021 | 4192 | 1362 | 5554 |
| Madagascar 2021 | 4580 | 812 | 5392 |
| Uganda 2016 | 3539 | 885 | 4424 |
| Mali 2018 | 3415 | 874 | 4289 |
| Cote d'Ivoire 2021 | 2259 | 2007 | 4266 |
| Mozambique 2022–23 | 2733 | 1043 | 3776 |
| Ghana 2022–23 | 1873 | 1795 | 3668 |
| Guinea 2012 | 2339 | 792 | 3131 |
| Togo 2013–14 | 1913 | 981 | 2894 |

**Supplementary Figure 1:** Percentage of malaria positivity in children aged 6-59 months tested for malaria by household occupation category, in urban and rural clusters combined.

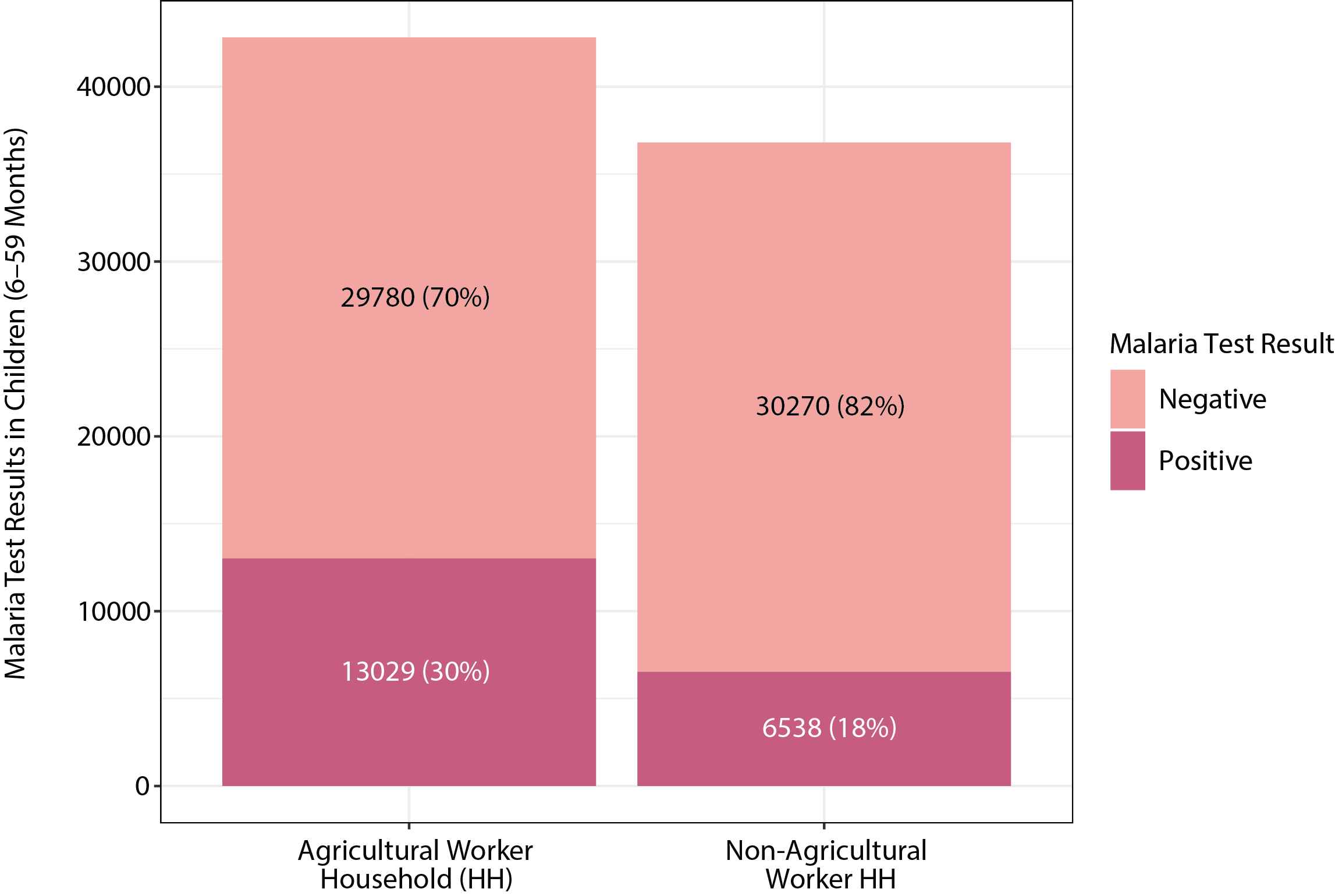

**Supplementary Figure 2.** Percentage of malaria positivity in children aged 6–59 months tested for malaria in urban and rural clusters per country.

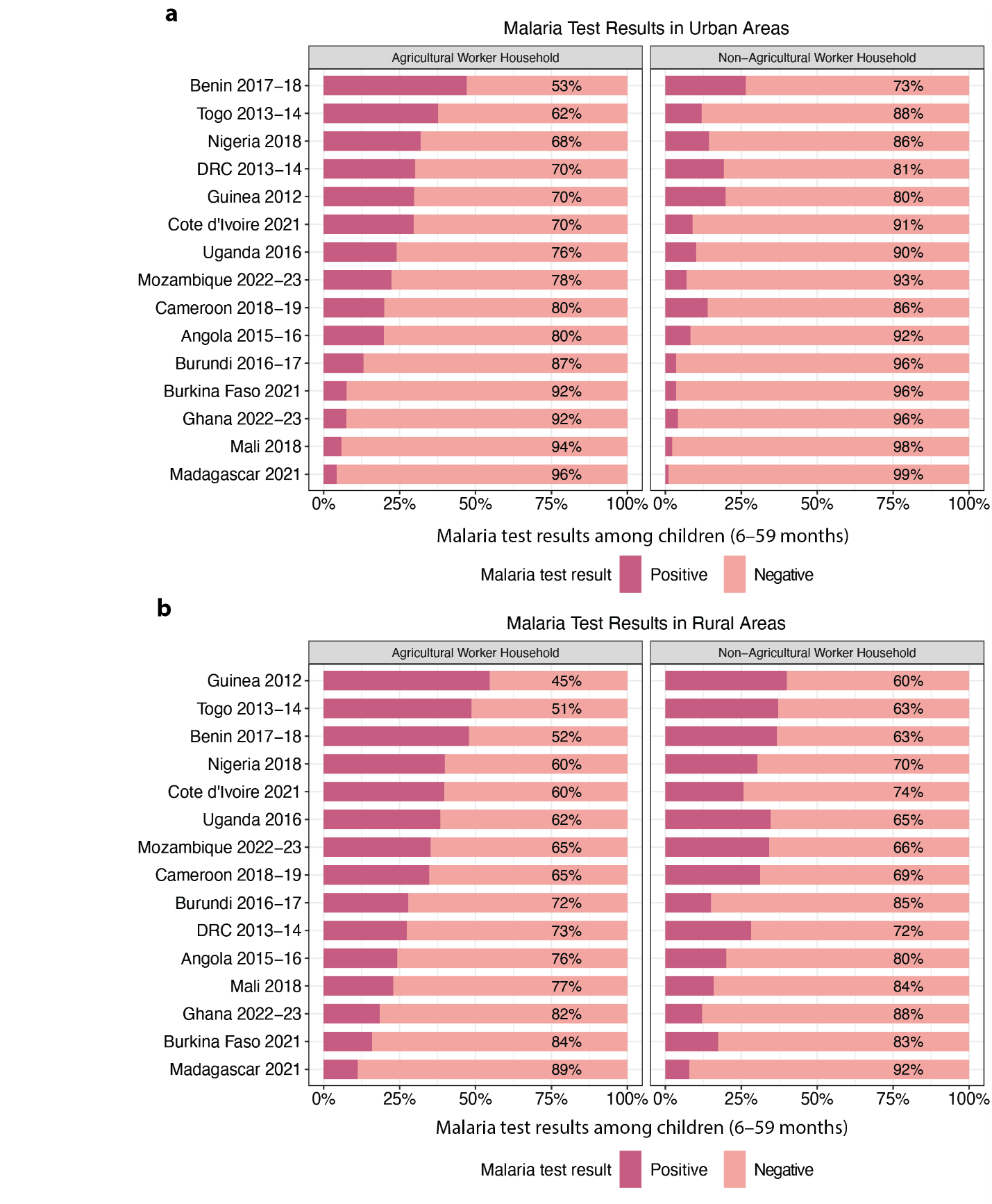

Supplementary Figure 2: Malaria test results in children aged 6–59 months who were tested for malaria in urban and rural clusters per country, stratified by household occupation category.

**Supplementary Table 2.** Adjusted Association of Covariates with Odds of Testing Positive for Malaria in Urban and Rural Populations. Model adjusts for home type (agricultural vs. non-agricultural). Table only includes variables considered for mediation.

| Covariate | Odds of testing positive for malaria (CI), Urban | Odds of testing positive for malaria (CI),  Rural |
| --- | --- | --- |
| Enhanced vegetation index | 1.483 (1.413 – 1.556) | 1.132 (1.096 – 1.169) |
| Housing quality |  |  |
| Non-modern house | 1.000 | 1.000 |
| Modern house | 0.581 (0.511 – 0.660) | 0.783 (0.724 – 0.847) |
| Household size | 1.027 (1.013 – 1.041) | 1.034 (1.026 – 1.042) |
| Prior night net use among children under five |  |  |
| Did not use | 1.000 | 1.000 |
| Used | 1.026 (0.922 – 1.141) | 0.820 (0.772 – 0.871) |
| Stunting |  |  |
| Not stunted | 1.000 | 1.000 |
| Stunted | 1.613 (1.392 – 1.870) | 1.447 (1.354 – 1.546) |
| Wealth index |  |  |
| Poorest | 1.000 | 1.000 |
| Poor | 0.525 (0.415 – 0.663) | 0.874 (0.813 – 0.940) |
| Middle | 0.394 (0.316 – 0.493) | 0.659 (0.607 – 0.715) |
| Rich | 0.290 (0.231 – 0.364) | 0.464 (0.415 – 0.519) |
| Richest | 0.135 (0.107 – 0.171) | 0.209 (0.171 – 0.254) |
| Treatment-Seeking |  |  |
| Less than or equal to 60% sought treatment | 1.000 | 1.000 |
| More than 60% sought treatment | 0.714 (0.619 – 0.823) | 0.866 (0.795 – 0.944) |

**Supplementary Table 3.** Unadjusted Association of Home Type with Covariates in Urban and Rural Populations. Table only includes variables considered for mediation.

| Term | Covariate (Dependent Variable) | Estimate Type* | Urban | Rural |
| --- | --- | --- | --- | --- |
| Home type: agricultural | Enhanced vegetation index | Coef | 0.962 (0.855 – 1.070) | 0.257 (0.193 – 0.321) |
| Home type: agricultural | Housing quality: modern | OR | 0.261 (0.229 – 0.298) | 0.330 (0.302 – 0.360) |
| Home type: agricultural | Household size | Coef | 1.303 (1.038 – 1.567) | 0.963 (0.814 – 1.111) |
| Home type: agricultural | Net use among children under five: used | OR | 1.268 (1.137 – 1.413) | 0.911 (0.849 – 0.977) |
| Home type: agricultural | Stunting: stunted | OR | 1.749 (1.494 – 2.047) | 1.368 (1.268 – 1.476) |
| Home type: agricultural | Wealth index | OR | 0.067 (0.052 – 0.086) | 0.535 (0.487 – 0.588) |
| Home type: agricultural | Treatment-seeking: Over 60% Sought | OR | 0.261 (0.229 – 0.298) | 0.330 (0.302 – 0.360) |

*Regression estimates are presented as coefficients (Coef) for continuous variables and odds ratios (OR) for binary or categorical variables. The independent variable in all models is home type (agricultural vs. non-agricultural), with non-agricultural as the reference category. Dependent variables are modeled individually; for binary outcomes, the reference category is the absence of the characteristic (e.g., not stunted, not modern housing).

**Supplementary Table 4.** Mediation Analysis Results

### **Urban**

| Mediator | Effect Type | Estimate | SE | Lower 95% CI | Upper 95% CI | P-Value | % Mediation | Bootstrapped Lower CI | Bootstrapped Upper CI |
| --- | --- | --- | --- | --- | --- | --- | --- | --- | --- |
| Stunting | Indirect | 0.0037 | 0.0005 | 0.0027 | 0.0047 | p < 0.0001 | 2.6994 | 1.8777 | 3.5740 |
| Stunting | Direct | 0.1350 | 0.0053 | 0.1246 | 0.1454 | p < 0.0001 | NA | NA | NA |
| Stunting | Total | 0.1387 | 0.0053 | 0.1283 | 0.1491 | p < 0.0001 | NA | NA | NA |
| Household Size | Indirect | 0.0043 | 0.0008 | 0.0028 | 0.0059 | p < 0.0001 | 3.1943 | 1.9880 | 4.4953 |
| Household Size | Direct | 0.1333 | 0.0053 | 0.1229 | 0.1437 | p < 0.0001 | NA | NA | NA |
| Household Size | Total | 0.1377 | 0.0053 | 0.1274 | 0.1480 | p < 0.0001 | NA | NA | NA |
| Wealth | Indirect | 0.0637 | 0.0024 | 0.0590 | 0.0684 | p < 0.0001 | 46.4459 | 41.7435 | 51.7795 |
| Wealth | Direct | 0.0739 | 0.0056 | 0.0629 | 0.0850 | p < 0.0001 | NA | NA | NA |
| Wealth | Total | 0.1377 | 0.0053 | 0.1274 | 0.1480 | p < 0.0001 | NA | NA | NA |
| Housing Quality | Indirect | 0.0193 | 0.0014 | 0.0165 | 0.0221 | p < 0.0001 | 14.0577 | 11.7884 | 16.5194 |
| Housing Quality | Direct | 0.1184 | 0.0054 | 0.1078 | 0.1290 | p < 0.0001 | NA | NA | NA |
| Housing Quality | Total | 0.1377 | 0.0053 | 0.1274 | 0.1480 | p < 0.0001 | NA | NA | NA |
| Net Use | Indirect | -0.0001 | 0.0001 | -0.0004 | 0.0001 | 0.3704 | -0.0926 | -0.3282 | 0.1015 |
| Net Use | Direct | 0.1378 | 0.0053 | 0.1275 | 0.1481 | p < 0.0001 | NA | NA | NA |
| Net Use | Total | 0.1377 | 0.0053 | 0.1274 | 0.1480 | p < 0.0001 | NA | NA | NA |
| EVI | Indirect | 0.0374 | 0.0016 | 0.0343 | 0.0406 | p < 0.0001 | 27.4059 | 24.3778 | 30.6996 |
| EVI | Direct | 0.0993 | 0.0054 | 0.0887 | 0.1098 | p < 0.0001 | NA | NA | NA |
| EVI | Total | 0.1367 | 0.0053 | 0.1263 | 0.1471 | p < 0.0001 | NA | NA | NA |
| Treatment-Seeking | Indirect | 0.0193 | 0.0014 | 0.0165 | 0.0221 | p < 0.0001 | 13.9904 | 11.6532 | 16.5099 |
| Treatment-Seeking | Direct | 0.1184 | 0.0054 | 0.1078 | 0.1290 | p < 0.0001 | NA | NA | NA |
| Treatment-Seeking | Total | 0.1377 | 0.0053 | 0.1274 | 0.1480 | p < 0.0001 | NA | NA | NA |

### **Rural**

| Mediator | Effect Type | Estimate | SE | Lower 95% CI | Upper 95% CI | P-Value | % Mediation | Bootstrapped Lower CI | Bootstrapped Upper CI |
| --- | --- | --- | --- | --- | --- | --- | --- | --- | --- |
| Stunting | Indirect | 0.0037 | 0.0004 | 0.0030 | 0.0044 | p < 0.0001 | 7.0099 | 5.3908 | 8.9807 |
| Stunting | Direct | 0.0491 | 0.0043 | 0.0406 | 0.0576 | p < 0.0001 | NA | NA | NA |
| Stunting | Total | 0.0528 | 0.0043 | 0.0443 | 0.0613 | p < 0.0001 | NA | NA | NA |
| Household Size | Indirect | 0.0056 | 0.0006 | 0.0045 | 0.0067 | p < 0.0001 | 10.7367 | 8.3703 | 13.4419 |
| Household Size | Direct | 0.0474 | 0.0043 | 0.0389 | 0.0559 | p < 0.0001 | NA | NA | NA |
| Household Size | Total | 0.0530 | 0.0043 | 0.0446 | 0.0615 | p < 0.0001 | NA | NA | NA |
| Wealth | Indirect | 0.0282 | 0.0011 | 0.0261 | 0.0304 | p < 0.0001 | 53.6051 | 45.2476 | 64.1169 |
| Wealth | Direct | 0.0248 | 0.0044 | 0.0163 | 0.0333 | p < 0.0001 | NA | NA | NA |
| Wealth | Total | 0.0530 | 0.0043 | 0.0446 | 0.0615 | p < 0.0001 | NA | NA | NA |
| Housing Quality | Indirect | 0.0094 | 0.0010 | 0.0075 | 0.0113 | p < 0.0001 | 17.7556 | 13.4862 | 22.8902 |
| Housing Quality | Direct | 0.0437 | 0.0044 | 0.0351 | 0.0523 | p < 0.0001 | NA | NA | NA |
| Housing Quality | Total | 0.0531 | 0.0043 | 0.0446 | 0.0615 | p < 0.0001 | NA | NA | NA |
| Net Use | Indirect | 0.0010 | 0.0002 | 0.0006 | 0.0015 | p < 0.0001 | 1.9966 | 1.2620 | 2.8858 |
| Net Use | Direct | 0.0520 | 0.0043 | 0.0436 | 0.0604 | p < 0.0001 | NA | NA | NA |
| Net Use | Total | 0.0530 | 0.0043 | 0.0446 | 0.0615 | p < 0.0001 | NA | NA | NA |
| EVI | Indirect | 0.0073 | 0.0005 | 0.0063 | 0.0084 | p < 0.0001 | 13.9346 | 11.2977 | 16.9912 |
| EVI | Direct | 0.0457 | 0.0043 | 0.0372 | 0.0541 | p < 0.0001 | NA | NA | NA |
| EVI | Total | 0.0530 | 0.0043 | 0.0445 | 0.0615 | p < 0.0001 | NA | NA | NA |
| Treatment-Seeking | Indirect | 0.0094 | 0.0010 | 0.0075 | 0.0113 | p < 0.0001 | 17.7842 | 13.1598 | 22.7262 |
| Treatment-Seeking | Direct | 0.0437 | 0.0044 | 0.0351 | 0.0523 | p < 0.0001 | NA | NA | NA |
| Treatment-Seeking | Total | 0.0531 | 0.0043 | 0.0446 | 0.0615 | p < 0.0001 | NA | NA | NA |

**Supplementary Table 5.** Environmental Variable Likelihood Ratio Test Results

(A) Urban

| **Variable name** | **Likelihood ratio test p-value** |
| --- | --- |
| Enhanced vegetation index | 8.4775e-15 |
| Precipitation (mm) | 0.00050868 |
| Relative humidity | 3.8212e-15 |
| Temperature | 0.93006 |

(B) Rural

| **Variable name** | **Likelihood ratio test p-value** |
| --- | --- |
| Enhanced vegetation index | 2.193e-06 |
| Precipitation (mm) | 0.52002 |
| Relative humidity | 0.064546 |
| Temperature | 0.016866 |

**Supplementary Table 6:** Population Estimates for 3 Most Populous First-Level Administrative Subdivisions

| **Country** | **Subdivision Name** | **Subdivision Type** | **Population Estimate** |
| --- | --- | --- | --- |
| Angola | Benguela | Province | 2009036 |
| Angola | Huíla | Province | 2609916 |
| Angola | Luanda | Province | 8732536 |
| Burkina Faso | Centre | Region | 4219205 |
| Burkina Faso | Est | Region | 2029244 |
| Burkina Faso | Haut-Bassins | Region | 2479624 |
| Benin | Atlantique | Department | 1688688 |
| Benin | Borgou | Department | 1534438 |
| Benin | Ouémé | Department | 1305924 |
| Burundi | Gitega | Province | 844019 |
| Burundi | Kirundo | Province | 792388 |
| Burundi | Muyinga | Province | 915425 |
| Democratic Republic of the Congo | Haut-Katanga | Province | 6332890 |
| Democratic Republic of the Congo | Kinshasa | Province | 6847929 |
| Democratic Republic of the Congo | Nord-Kivu | Province | 7196505 |
| Côte d'Ivoire | Abidjan | District | 5477720 |
| Côte d'Ivoire | Montagnes | District | 2868437 |
| Côte d'Ivoire | Sassandra-Marahoué | District | 2763779 |
| Cameroon | Centre | Region | 5544560 |
| Cameroon | Extrême-Nord | Region | 4521621 |
| Cameroon | Littoral | Region | 4259234 |
| Ghana | Ashanti | Region | 6759214 |
| Ghana | Eastern | Region | 3355397 |
| Ghana | Greater Accra | Region | 5632346 |
| Guinea | Conakry | Region | 1570273 |
| Guinea | Kankan | Region | 1823141 |
| Guinea | Nzérékoré | Region | 1540905 |
| Madagascar | Analamanga | Region | 4713540 |
| Madagascar | Vakinankaratra | Region | 1877858 |
| Madagascar | Vatovavy Fitovinany | Region | 1896047 |
| Mali | Koulikoro | Region | 3950045 |
| Mali | Ségou | Region | 3135600 |
| Mali | Sikasso | Region | 3763009 |
| Mozambique | Nampula | Province | 6047604 |
| Mozambique | Tete | Province | 3535341 |
| Mozambique | Zambezia | Province | 5785796 |
| Nigeria | Kaduna | State | 8695951 |
| Nigeria | Kano | State | 13815361 |
| Nigeria | Lagos | State | 13207366 |
| Togo | Maritime | Region | 2917236 |
| Togo | Plateaux | Region | 1521410 |
| Togo | Savanes | Region | 950788 |
| Uganda | Central | Region | 10243258 |
| Uganda | Eastern | Region | 9684526 |
| Uganda | Western | Region | 9481493 |

**Supplementary Figure 3.** Percentages of prior night net use in children aged 6–59 months who were tested for malaria in urban and rural clusters per count­ry.

**(a**) Urban

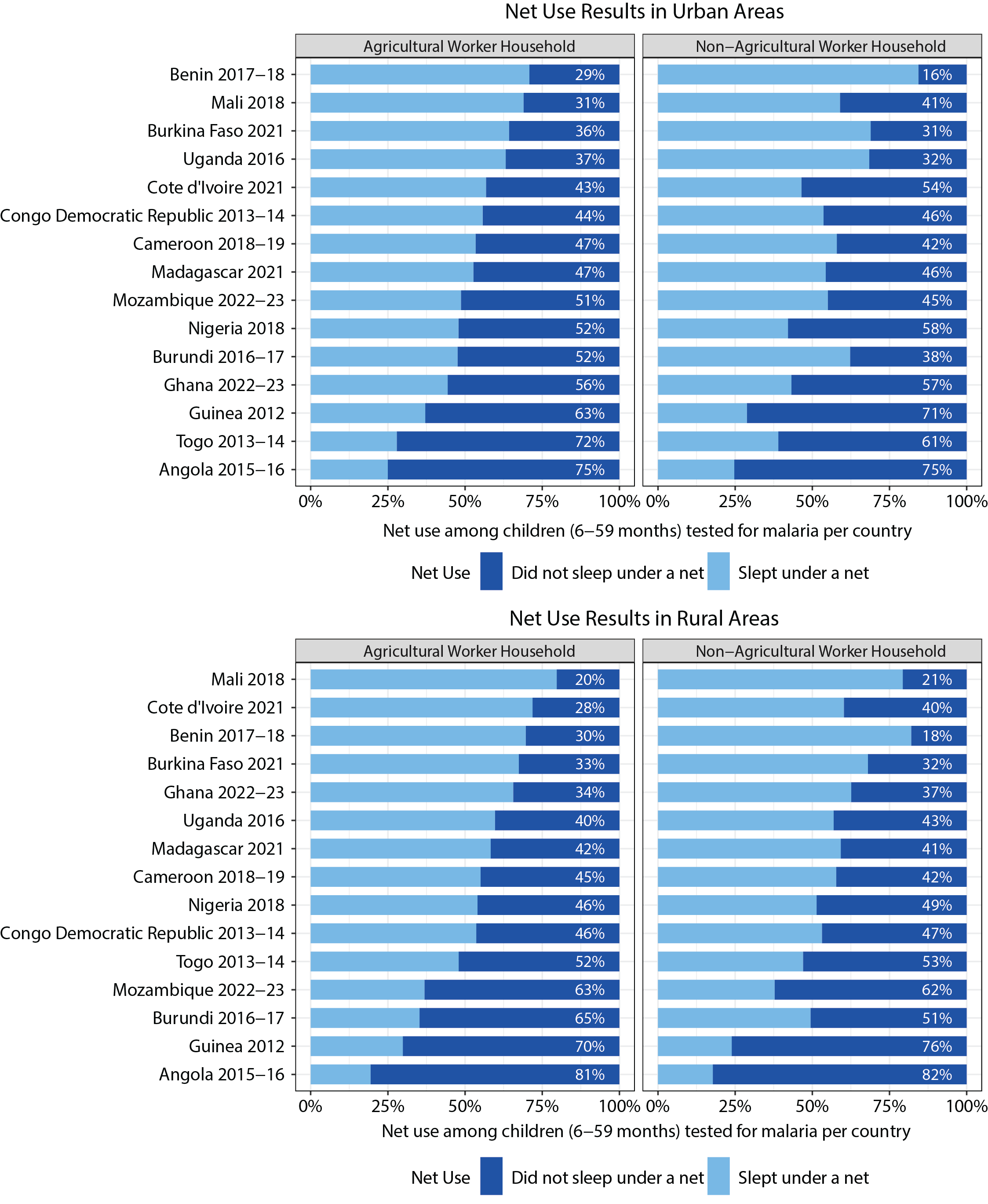

**(b**) Rural

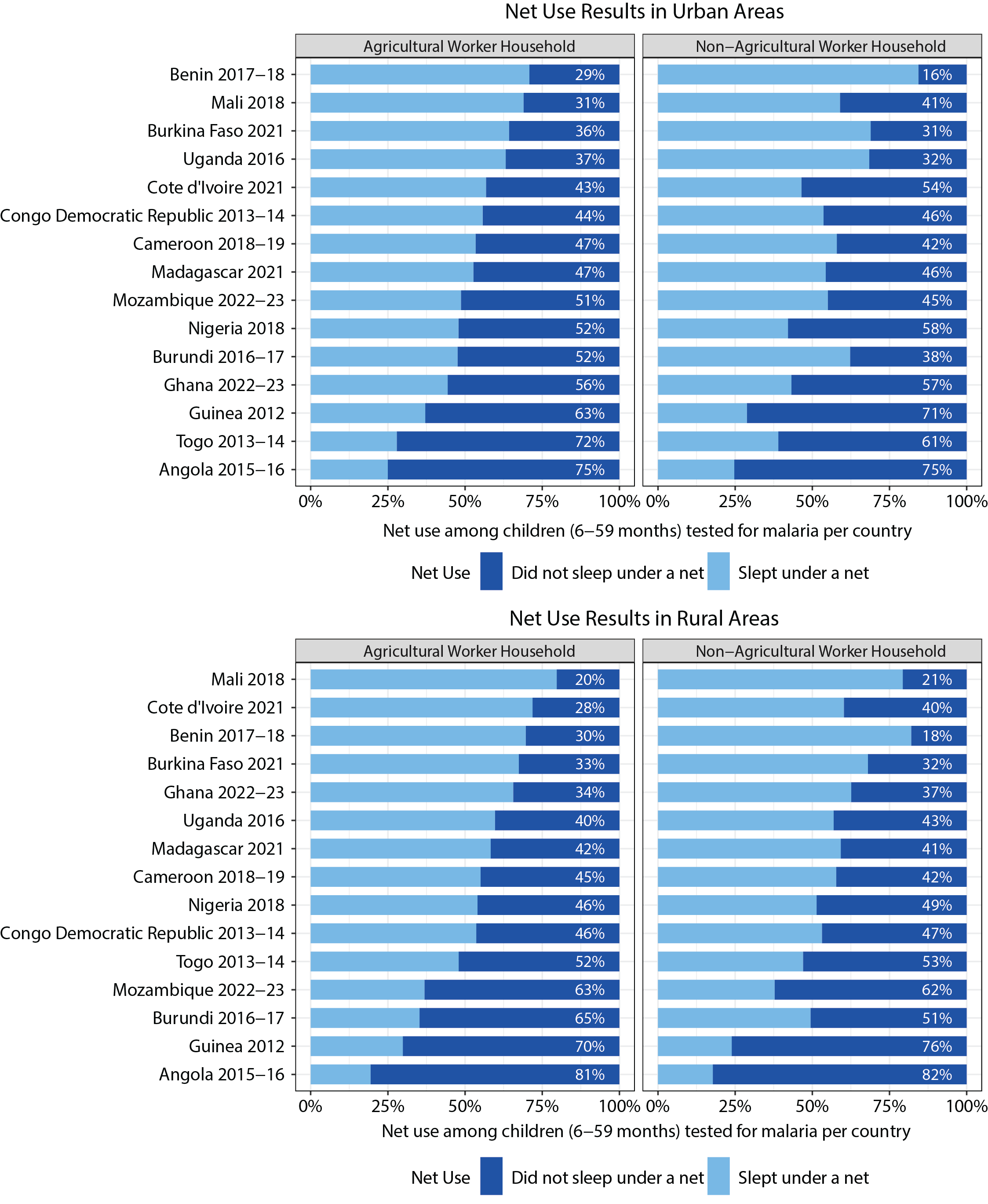

Supplementary Figure 3: Percentages of net use the night prior to the survey in children aged 6–59 months who were tested for malaria in urban and rural clusters per country, stratified by household occupation category.

**Supplementary Figure 4**. Malaria test positivity rate and net use rate in children under 5 with percentage change in number of nets distributed yearly compared to first year of distribution.

**(a)** Urban

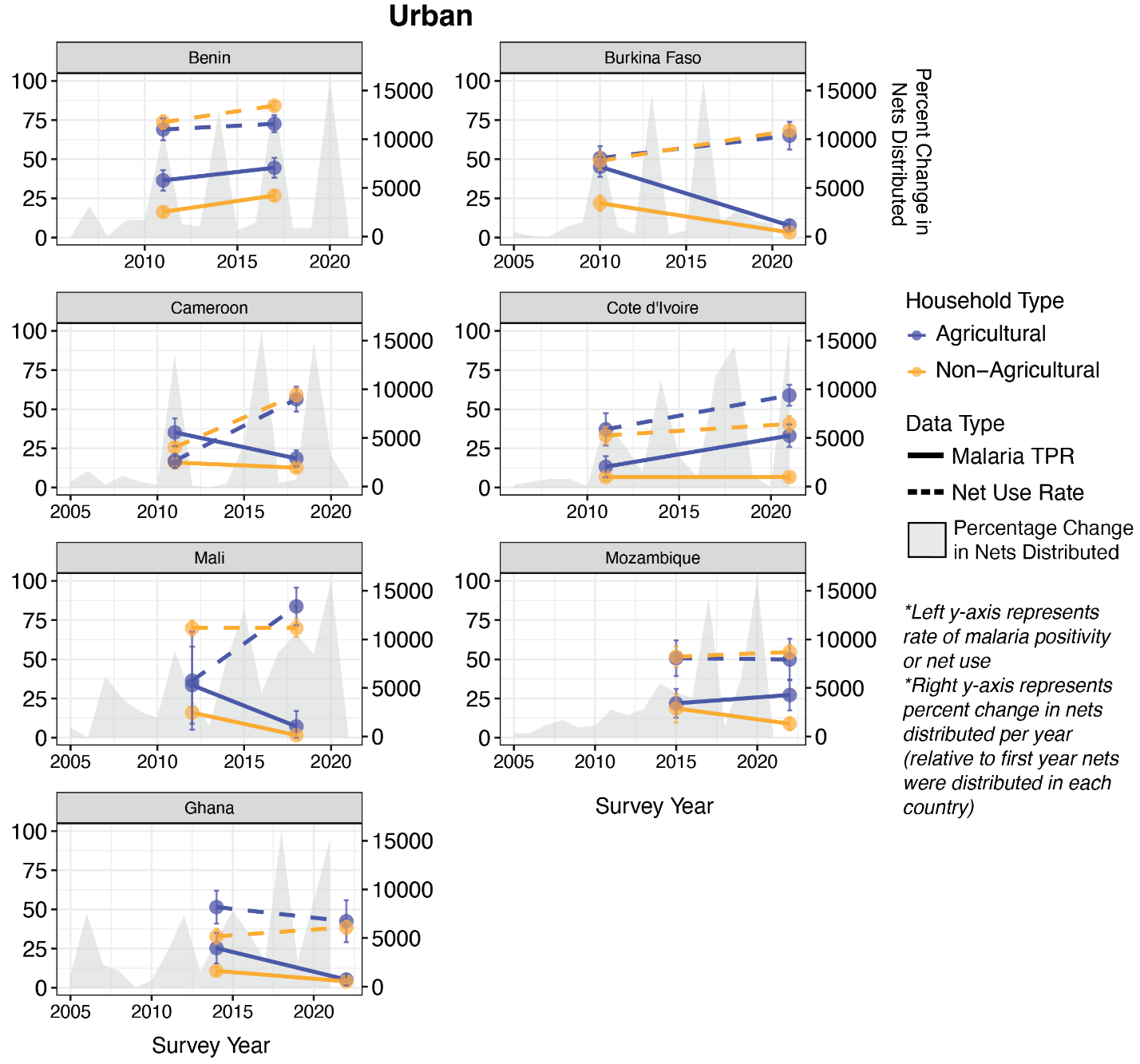

**(b)** Rural

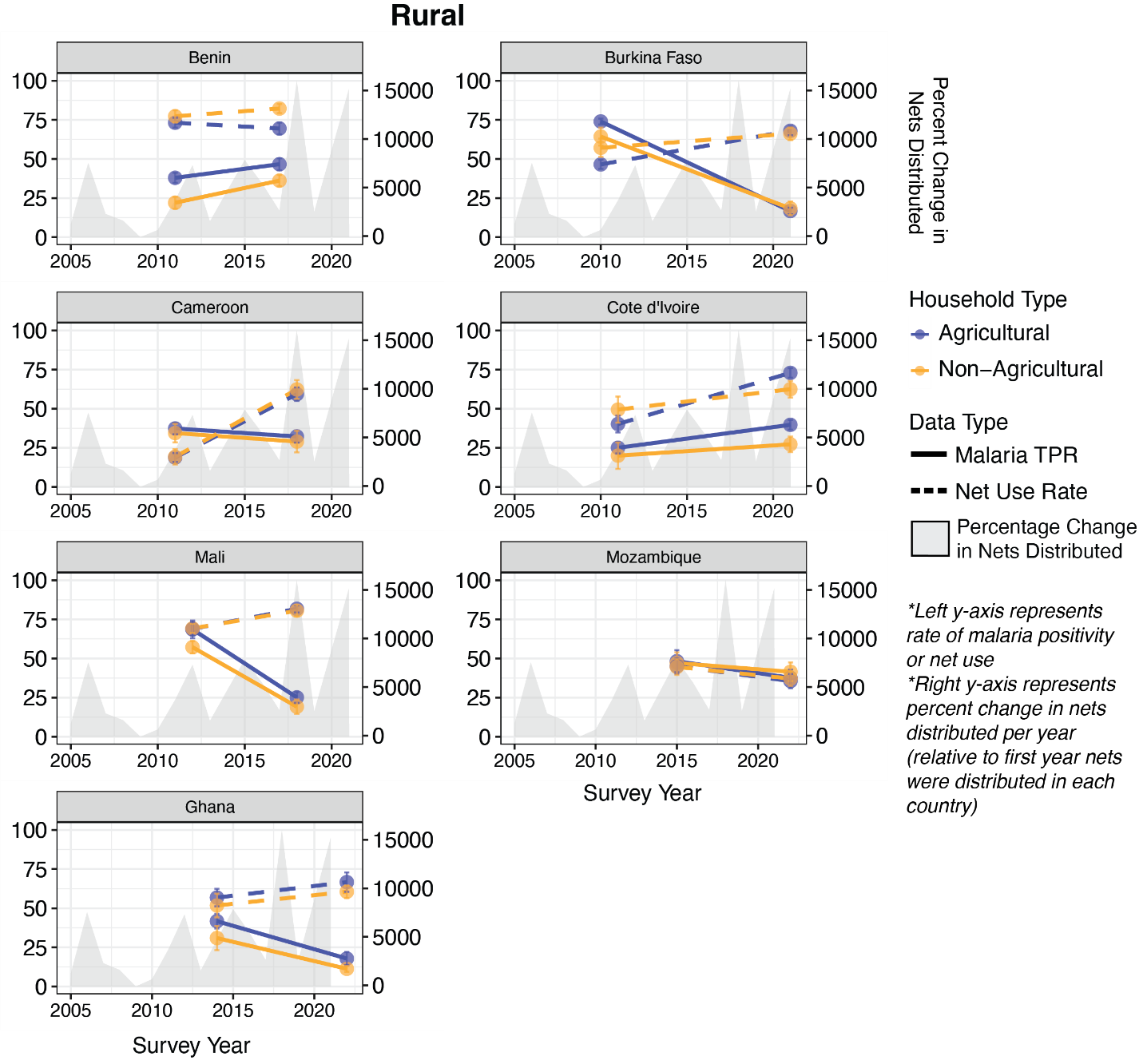

Supplementary Figure 4: **(a)** Trends in test positivity rates and net use rates between agricultural worker HHs and non-agricultural worker HHs within urban clusters per country with both preceding and recent survey data included in the analysis. **(b)** Trends in test positivity rates and net use rates between agricultural worker HHs and non-agricultural worker HHs within rural clusters per country with both preceding and recent survey data included in the analysis. The left y-axis on each plot represents the rate of malaria positivity or net use, while the right y-axis represents the percent change in nets distributed per year relative to the first year nets were distributed in the given country. The grey area plot shows this percentage change in nets distributed per year.

**Supplementary Figure 5.** Percent Mediated by Key Mediators Across Countries, Stratified by Urban and Rural Residence

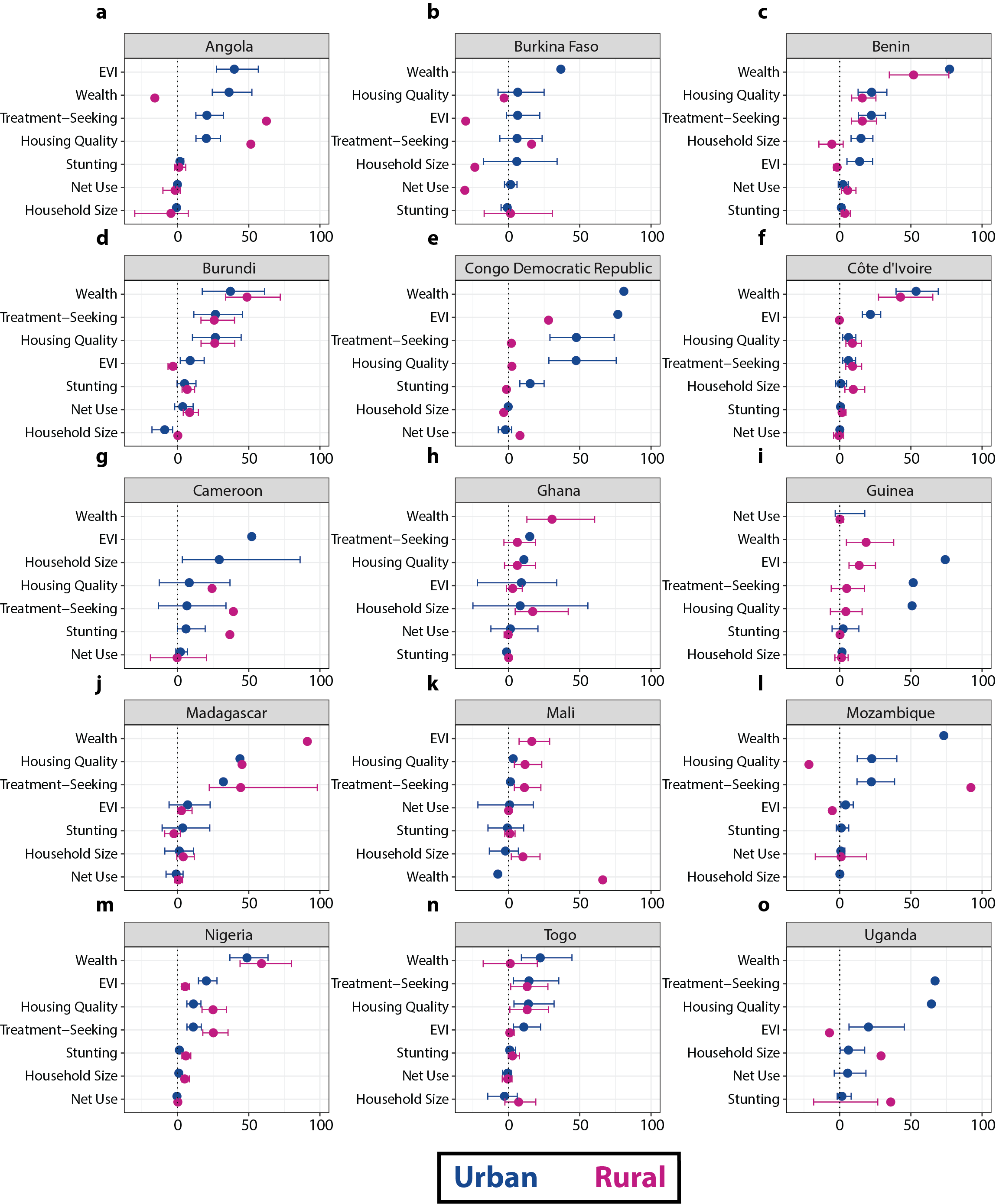

Supplementary Figure 5: **(a–o)** Percent mediated by various mediators of the relationship between home type and malaria test positivity rate, stratified by country and by urban and rural place of residence. Error bars represent 95% confidence intervals.

**Supplementary Figure 6.** Odds ratios and predicted probabilities of testing positive for malaria (by RDT or microscopy) in children aged 6–59 months from urban and rural households combined by home type and after adjustment with each potential mediator.

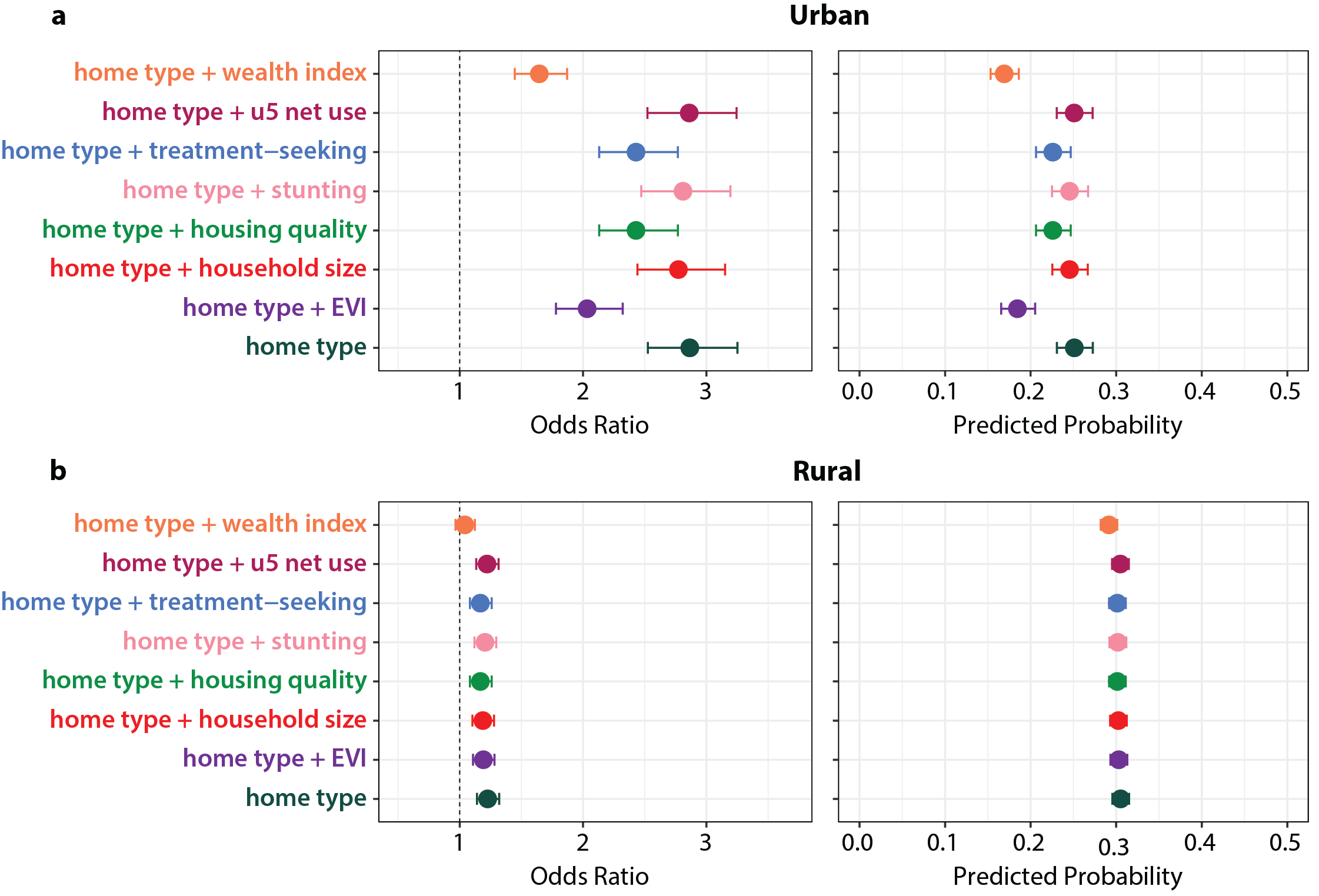

**Supplementary Figure 7.** Mediation Analysis Results (Indirect, Direct, and Total Effects for each Mediator)

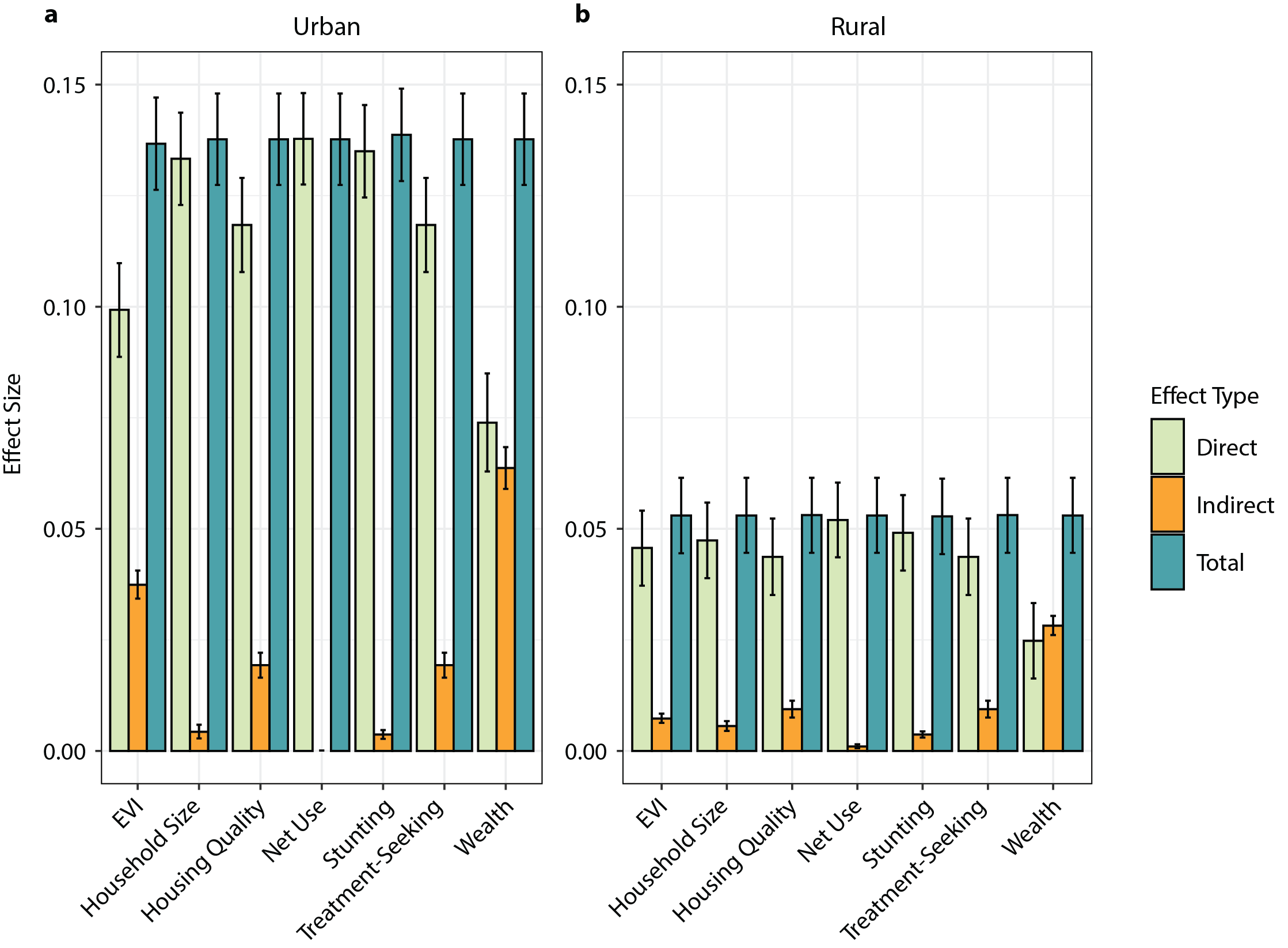

Supplementary Figure 7: Direct, indirect, and total effects calculated via mediation analysis for various mediators of the relationship between home type and malaria test positivity rate. Error bars represent 95% confidence intervals.

**Supplementary Figure 8.** Directed Acyclic Graph (DAG) Used to Model Potential Mediators

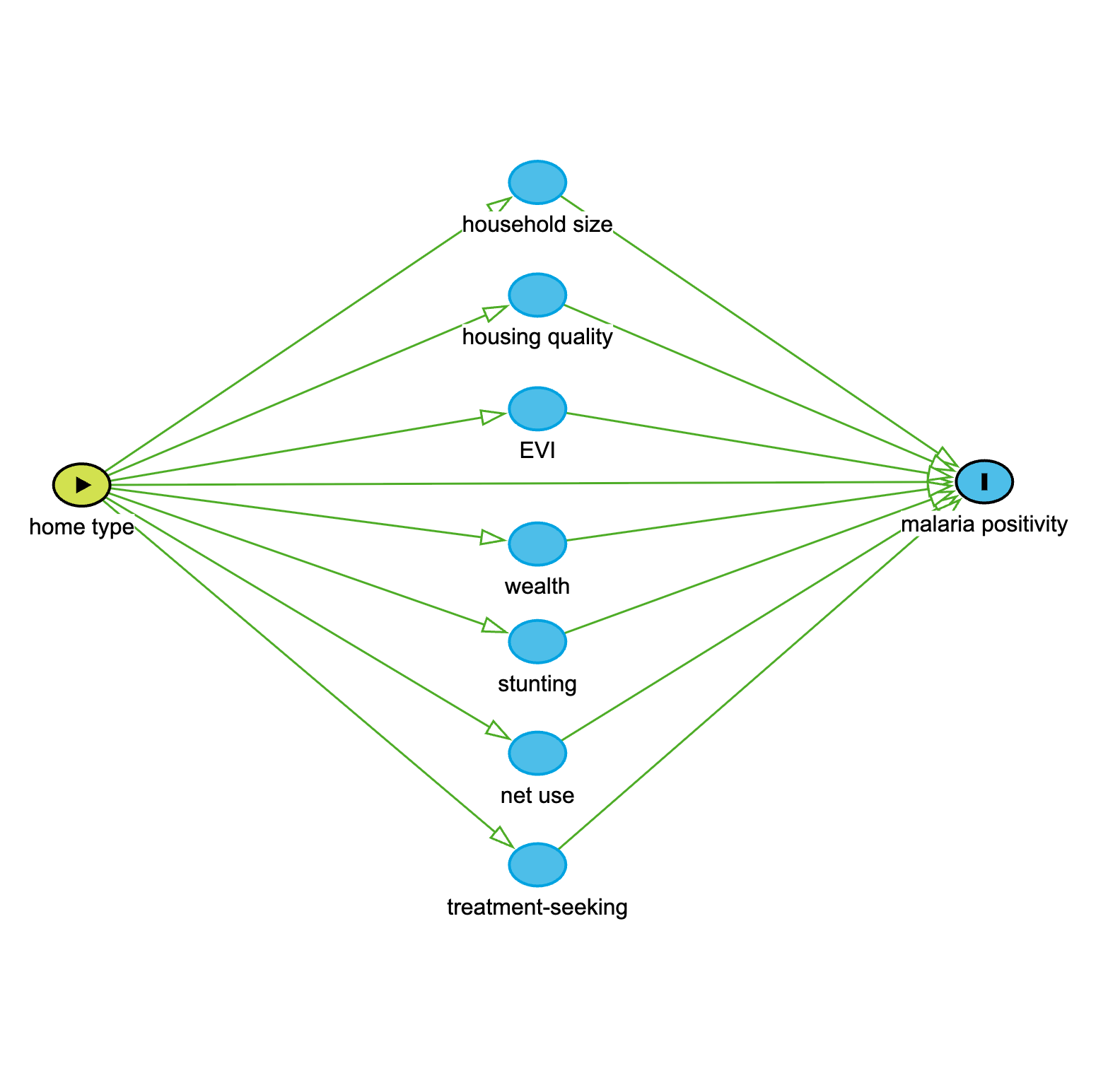
